# Incidence and Survival of Head and Neck Cancers in the United Kingdom 2000-2021

**DOI:** 10.1101/2024.12.05.24318538

**Authors:** Andrea Miquel Dominguez, Eng Hooi Tan, Edward Burn, Antonella Delmestri, Talita Duarte-Salles, Asieh Golozar, Wai Yi Man, Daniel Prieto-Alhambra, Francesc Xavier Avilés-Jurado, Danielle Newby

## Abstract

**Background:** Understanding about the changing burden of head and neck cancers (HNC) is essential to guide public health interventions and inform cancer care strategies.

**Methods:** We conducted a population-based cohort study using routinely collected primary care data Clinical Practice Research Datalink (CPRD) GOLD from the United Kingdom. Adults aged ≥18 years with ≥1 year of prior history were included. We estimated crude and age-standardised incidence rates (IRs) and one-, five-, and ten-year survival from 2000 to 2021, stratified by age and calendar year. Findings from CPRD GOLD were compared with primary care data from CPRD Aurum, which includes patients from England only.

**Results:** There were 12,455 patients with a diagnosis of HNC from CPRD GOLD (69.2% male; median age 64 years). Crude incidence in GOLD increased from 9.08 (95% CI: 7.88–10.42) per 100,000 person-years in 2000 to 15.59 (14.07–17.23) in 2021, with similar trends observed in CPRD Aurum. Age-standardised incidence trends were attenuated overall but remained elevated for oropharyngeal and tongue cancers. Five-year survival improved modestly, from 53.8% (95% CI: 51.4–56.3%) in 2000–2004 to 58.7% (56.5–60.9%) in 2015–2019.

**Conclusions:** HNC increases over recent decades are likely due to ageing with increases in oropharyngeal cancers likely due to changing behavioural risk factors. Small improvements in long term survival highlights more research is needed to improve earlier diagnosis which will lead to better patient outcomes.

## INTRODUCTION

Head and neck cancers (HNC) are a heterogenous group of cancers affecting the upper aerodigestive tract including the oral cavity, pharynx, larynx, paranasal sinuses, nasal cavity, and salivary glands(1,2). Combined, these cancers are the sixth most common cancer worldwide accounting for more than 940,000 new cases and 480,000 deaths in 2022(3).

The etiology and epidemiology of HNC subsites can vary with distinct characteristics in terms of histology subtypes, symptoms, treatment approaches, and prognosis, making it necessary to consider them as separate entities(4). The overall incidence of HNC has been shown to be increasing over time with marginal improvements in survival in the United Kingdom (UK) and in other countries(5) However, specific risk factors such as tobacco and alcohol consumption and human papillomavirus (HPV) infection have impacted the disease burden of the different HNC subtypes(6).

Understanding trends in incidence, and overall survival of HNC and subsites is an important aspect to inform decisions regarding screening, prevention, treatment, and disease management. Due to changes in HPV infections and vaccinations, a decline in tobacco and alcohol usage in recent years, an up-to-date comprehensive assessment of the trends of HNC and subsites is lacking. The aim of this study is to describe HNC and subsite trends in terms of incidence and survival from 2000-2021 using primary care data from the UK.

## MATERIALS AND METHODS

### Study design, setting, and data sources

We carried out a population cohort study using routinely collected primary care data from the UK. People with a diagnosis of HNC and a denominator population were identified from the CPRD GOLD database. We repeated the study using CPRD Aurum for comparison. CPRD GOLD includes data from practices using the Vision® system across the UK (England, Wales, Northern Ireland and Scotland) and has been cited to represent between 4.3 - 6.9% of the UK population whereas CPRD Aurum includes data from practices using the EMIS Web® system in England only and represents 13% of the population in 2018(7–9). Both databases contain pseudonymised patient-level information on demographics, lifestyle data, clinical diagnoses, prescriptions, and preventive care and are broadly representative. In recent years, the regional distribution of currently contributing practices for GOLD has significantly shifted, resulting in many new practices joining from Scotland, Wales, and Northern Ireland, and fewer participating from England. Therefore, the justification to include both databases is to allow for comparison across the whole of the UK. Both databases were mapped to the Observational Medical Outcomes Partnership Common Data Model(10).

### Study participants

Eligible individuals were 18 years or older with one year of prior history, in the database from 1st January 2000. These individuals were followed up to whichever came first: HNC diagnosis, exit from database, date of death, or study end (December 31, 2021, for GOLD, and, due to data availability December 31, 2019, for Aurum). For survival analysis, individuals were followed from HNC diagnosis to either death, exit from database, or study end.

### Outcome definitions

We used diagnostic clinical codes to identify HNC and subsites. Subsites of HNC were categorised as cancers of the oral cavity, tongue and lingual tonsils, nasal cavity and sinuses, salivary glands, hypopharynx, nasopharynx, oropharynx, and larynx.

Due to limitations in the specificity of diagnostic coding within primary care records, we grouped all tongue cancers into a single subsite. Although the tongue is anatomically divided into the anterior two-thirds (oral cavity) and the base (oropharynx), this distinction is often not captured, where non-specific codes such as “Malignant neoplasm of tongue” are commonly used. To address this, we created a dedicated codelist for cancers of the tongue to allowing inclusion of cases that would otherwise be excluded due to ambiguous classification.

Clinical codelists are provided in supplement with all analytical code in GitHub to enable reproducibility(11). For survival analyses, mortality was defined as all-cause mortality.

### Statistical methods

The characteristics of HNC patients were summarised, with median (IQR) for continuous variables and counts with percentages for categorical variables.

For incidence, number of events, observed time at risk, and incidence rates (IR) per 100,000 person years were summarised along with 95% confidence intervals (CI). Age-standardized IR were calculated using the 2013 European Standard Population(12). Age-standardized IR were compared to national cancer registry statistics across the UK(13–16).

For overall survival, we used the Kaplan-Meier method to estimate median survival and survival one, five, and ten years after diagnosis. Patients whose death and cancer diagnosis occurred on the same date were removed.

All results were stratified by sex and by age group. Survival estimates were additionally stratified by calendar time of cancer diagnosis in GOLD only. To avoid possible patient re-identification, results with fewer than five cases were suppressed.

Analyses were conducted using R version 4.2.3, with the *IncidencePrevalence(17)* and *survival* (18) packages. As this was a descriptive study, no formal statistical testing was conducted to assess temporal trends.

## RESULTS

### Patient characteristics

Overall, there were 11,388,117 eligible patients, with at least one year of prior history from 2000-2021 in GOLD with population attrition in the supplement (Supplement Table 2). Baseline patient characteristics of HNC patients is shown in Table 1.

**Table 1:** Demographics of HNC patients at the time of diagnosis for CPRD GOLD.

| Database | CPRD GOLD |
| --- | --- |
| Number of patients | 12455 |
| Sex: Male (N [%]) | 8614 (69.2%) |
| Age (years) (Median [IQR]) | 64 (56 to 73) |
| Age Groups N (years) (%) |  |
| 18-29 | 95 (0.8%) |
| 30-39 | 258 (2.1%) |
| 40-49 | 1071 (8.6%) |
| 50-59 | 3001 (24.1%) |
| 60-69 | 3730 (29.9%) |
| 70-79 | 2790 (22.4%) |
| 80-89 | 1293 (10.4%) |
| 90+ | 217 (1.7%) |
| Prior history, days |  |
| median [p25 - p75] | 3368 (1,818 to 5,119) |
| Smoking Status (5 years prior) |  |
| Non-smoker | 3187 (25.6%) |
| Former smoker | 92 (0.7%) |
| Current smoker | 5322 (42.7%) |
| Missing | 3854 (30.9%) |
| **General conditions (any time prior)** |  |
| Cerebrovascular disease | 727 (5.8%) |
| Chronic liver disease | 178 (1.4%) |
| Chronic obstructive lung disease | 1217 (9.8%) |
| Coronary arteriosclerosis | 115 (0.9%) |
| Dementia | 163 (1.3%) |
| Depressive disorder | 1655 (13.3%) |
| Diabetes mellitus | 1097 (8.8%) |
| Gastroesophageal reflux disease | 330 (2.6%) |
| Gastrointestinal haemorrhage | 797 (6.4%) |
| Heart disease | 1865 (15.0%) |
| Hyperlipidemia | 996 (8.0%) |
| Hypertensive disorder | 2966 (23.8%) |
| Ischemic heart disease | 1029 (8.3%) |
| Osteoarthritis | 1938 (15.6%) |
| Peripheral vascular disease | 379 (3.0%) |
| Renal impairment | 1067 (8.6%) |
| Venous thrombosis | 451 (3.6%) |

**Table 2:**
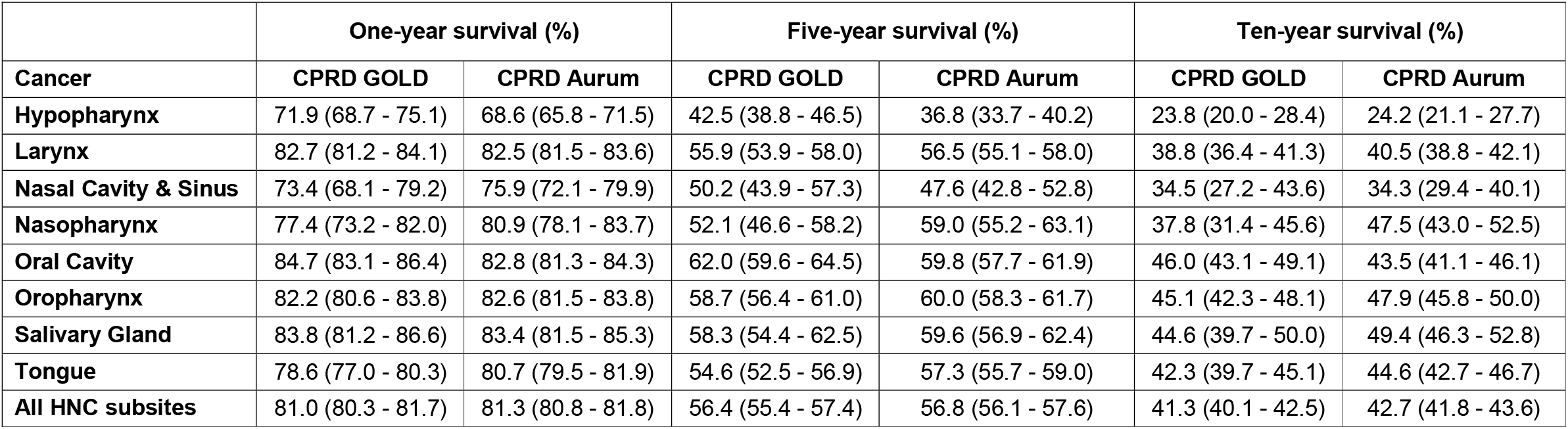
Survival rates (and 95% confidence intervals) of head and neck cancer subsites from in CPRD GOLD (2000-2021) and CPRD Aurum (2000-2019) stratified by database.

|  | One-year survival (%) |  | Five-year survival (%) |  | Ten-year survival (%) |  |
| --- | --- | --- | --- | --- | --- | --- |
| Cancer | CPRD GOLD | CPRD Aurum | CPRD GOLD | CPRD Aurum | CPRD GOLD | CPRD Aurum |
| Hypopharynx | 71.9 (68.7 - 75.1) | 68.6 (65.8 - 71.5) | 42.5 (38.8 - 46.5) | 36.8 (33.7 - 40.2) | 23.8 (20.0 - 28.4) | 24.2 (21.1 - 27.7) |
| Larynx | 82.7 (81.2 - 84.1) | 82.5 (81.5 - 83.6) | 55.9 (53.9 - 58.0) | 56.5 (55.1 - 58.0) | 38.8 (36.4 - 41.3) | 40.5 (38.8 - 42.1) |
| Nasal Cavity & Sinus | 73.4 (68.1 - 79.2) | 75.9 (72.1 - 79.9) | 50.2 (43.9 - 57.3) | 47.6 (42.8 - 52.8) | 34.5 (27.2 - 43.6) | 34.3 (29.4 - 40.1) |
| Nasopharynx | 77.4 (73.2 - 82.0) | 80.9 (78.1 - 83.7) | 52.1 (46.6 - 58.2) | 59.0 (55.2 - 63.1) | 37.8 (31.4 - 45.6) | 47.5 (43.0 - 52.5) |
| Oral Cavity | 84.7 (83.1 - 86.4) | 82.8 (81.3 - 84.3) | 62.0 (59.6 - 64.5) | 59.8 (57.7 - 61.9) | 46.0 (43.1 - 49.1) | 43.5 (41.1 - 46.1) |
| Oropharynx | 82.2 (80.6 - 83.8) | 82.6 (81.5 - 83.8) | 58.7 (56.4 - 61.0) | 60.0 (58.3 - 61.7) | 45.1 (42.3 - 48.1) | 47.9 (45.8 - 50.0) |
| Salivary Gland | 83.8 (81.2 - 86.6) | 83.4 (81.5 - 85.3) | 58.3 (54.4 - 62.5) | 59.6 (56.9 - 62.4) | 44.6 (39.7 - 50.0) | 49.4 (46.3 - 52.8) |
| Tongue | 78.6 (77.0 - 80.3) | 80.7 (79.5 - 81.9) | 54.6 (52.5 - 56.9) | 57.3 (55.7 - 59.0) | 42.3 (39.7 - 45.1) | 44.6 (42.7 - 46.7) |
| All HNC subsites | 81.0 (80.3 - 81.7) | 81.3 (80.8 - 81.8) | 56.4 (55.4 - 57.4) | 56.8 (56.1 - 57.6) | 41.3 (40.1 - 42.5) | 42.7 (41.8 - 43.6) |

There were 12,455 patients with HNC, 69% were male, with a median age of 64 years (IQR 56–73). The highest proportion of diagnoses occurred in the 60–69 age group (30%), with similar results in Aurum (Supplement Table 3).

By subsite (Supplement Table 4), the most common HNC were the larynx (22%), tongue (21%), and oropharynx (18.6%), while nasal cavity and sinus cancers were the least common (2.1%). Males accounted for most cases across all subsites, with the highest in laryngeal cancer (81%). The 60–69 age group represented the largest proportion of cases for most subsites, except for nasal cavity and sinus, and salivary gland cancers, where patients were older. In contrast, nasopharyngeal cancer was most frequently diagnosed in those aged 50–59. Patients with cancers of the hypopharynx, larynx and oral cavity had higher percentages of comorbidities. For most subsites (larynx, oral cavity, tongue, oropharynx, hypopharynx), smokers made up the highest proportion of patients (37% to 54%).

### Incidence rates stratified by calendar year, age, and sex

In GOLD, the crude overall IR of HNC from 2000 to 2021 was 14.2 (14.0 to 14.5) per 100,000 person-years. Males had higher overall IR (20.0 [19.6 to 20.4]) compared to females (8.6 [8.4 to 8.9]), with similar values in Aurum. Among subsites, laryngeal cancer had the highest IR (3.2 per 100,000 person-years) whereas nasal cavity and sinuses (0.3 per 100,000 for both databases) had the lowest. Across all subsites, males had higher overall IR compared to females (Supplement Table 5).

**Figure 1:**
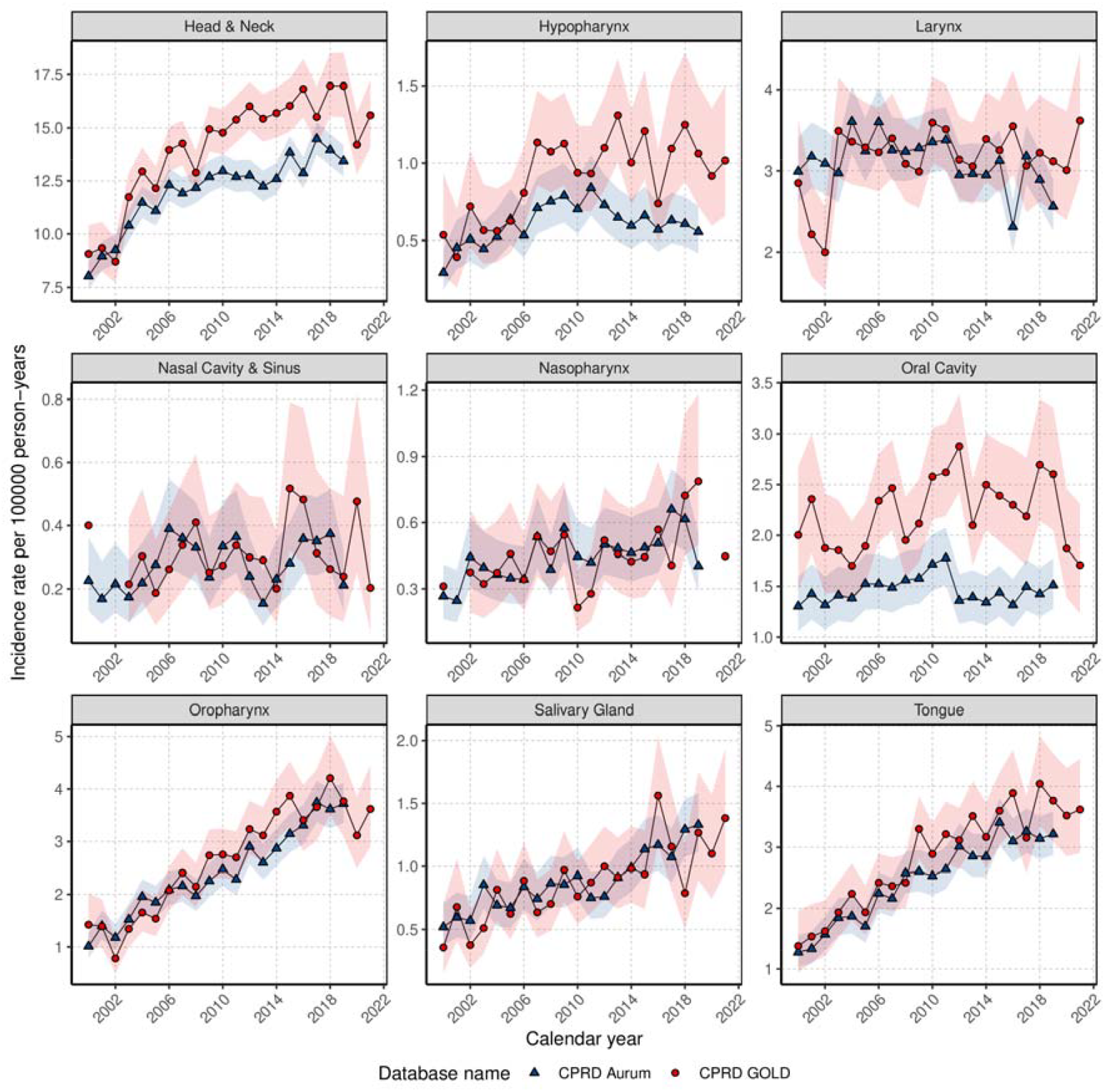
Crude annualised incidence rates for HNC and subsites stratified by database. Crude annualised IR for HNC increased across the study period (Figure 1). Subsite-specific trends showed rising IRs for cancers of the nasopharynx, oropharynx, salivary glands, and tongue. IR for hypopharyngeal cancer increased from 2000 to 2007 before stabilising, while IRs for laryngeal and nasal cavity and sinus cancers remained relatively stable. Oral cavity cancer showed a gradual increased from 2000 before dropping in 2012 and a subsequent rise. Similar trends were seen in Aurum except for hypopharyngeal cancer, which showed a gradual decline over time from 2007. Age standardized IR show trends have attenuated or stabilised except for tongue and oropharyngeal cancers, where increases are still seen, particularly in males (Supplement Figure 2). Comparison with national cancer registry statistics showed lower but comparable trends for overall HNC in GOLD (Supplement Figure 3).

Overall IR increased with age peaking at 70-79 years old: 33.2 (31.9 to 34.4) per 100,000 person-years in GOLD with similar values in Aurum (28.5 [27.7 to 29.3]) (Supplement Figure 4). Age specific rates showed stable trends in those aged 30-39 and 90+, while rates in those aged 40-59 and 80-89 stabilising after 2006. IR for those aged 60-79 gradually increased over time (Supplement Figure 5) with consistently higher rates in males (Supplement Figure 6).

Subsite-specific age trends showed similar patterns. Cancers of the hypopharynx and oral cavity, IR remained relatively stable for 50–59-year-olds. Laryngeal cancer showed declining IR in those aged 50-69 years in contrast to oropharyngeal, salivary gland, and tongue cancers, where IR increased for all age groups (Supplement Figure 7).

### Survival with age, sex, and calendar year stratification

In GOLD there were 12,381 patients with 5,662 deaths (46% of patients) over the study period with a median survival of 7.0 (6.6 - 7.5) years. Survival after one, five, and ten years after diagnosis was 81%, 56%, and 41% with similar results in Aurum (Supplement Figure 8). Females had a longer median survival (8.2 years) compared to males (6.5 years) with better five-year survival (59% Females 55%, Males). Cancer of the hypopharynx had the lowest median survival of 3.4 (2.9-4.1) years with cancers of the oral cavity showing the longest median survival of 8.7 [8.0 - 9.8]) years (Supplement Table 6).

Short, and long-term survival was similar for most subsites with one-, five- and ten-year survival estimated at 77-83%, 50-60% and 40-50% respectively apart from cancers of the hypopharynx and the nasal cavity and sinuses which were lower (Table 2). Stratification by calendar year (Supplement Figure 9) showed short-term survival has not improved but long-term survival has improved from 53.8% (51.4-56.3) in 2000-2004 to 58.7% (57-61) in 2015-2019. However, survival of oral cavity cancer decreased from 90% (87-93) in 2000-2004 to 83% (79-86) in 2015-2019 and nasopharyngeal cancer improved from 32.8% (22.0-49.0) to 63.1% (52.4-76.0) in five-year survival with wide confidence intervals.

## DISCUSSION

This study provides a comprehensive analysis of long-term trends in HNC incidence and survival in the UK. We observed notable increases in incidence, particularly among males and individuals aged 70–79, likely driven by an ageing population. However, increases were most pronounced for cancers of the tongue, salivary glands, and oropharynx, while laryngeal cancer incidence appeared stable or declining. Survival was poorest for hypopharyngeal and nasal cavity and sinus cancers. Although short-term outcomes remain unchanged, there has been a modest improvement in long-term survival.

Our findings align with existing trends, which have reported increases in HNC incidence over recent decades, particularly for cancers of the tongue, oropharynx, and salivary glands(5,19). While our IR broadly align with National Cancer Statistics across the UK, variations across countries, particularly for specific subsites, are potentially due to different classification of subsites as well as differing risk factors among the underlying populations(20). However, evidence suggests demographic profiles have remained unchanged over time despite rising oropharyngeal cancer incidence and stable or declining laryngeal cancer rates(21).

The reasons for increasing HNC cases are multifaceted and vary per subsite due to different etiologies and risk factors(20). Smoking and alcohol consumption are well-known behavioral risk factors, particularly among males, and contribute notably to oral cavity and laryngeal cancers(1,22). However, declines in cases in countries with higher sociodemographic indexes have been observed(23) with overall decline in these behaviors in the UK(24,25). This suggests rises in HNC cannot be attributed solely to smoking and alcohol consumption.

The rising incidence of HNC may be partly driven by increases in HPV infections(26) ^31^ which has led to notable increases in HPV-associated HNC, with rates for most non-HPV-related HNC remaining stable or declining(27). However, despite a potential link between HPV and HNC subsites(28), some evidence suggests that despite a doubling in oropharyngeal cancer cases, the proportion of cases attributable to HPV has not changed significantly, indicating that HPV likely contributes to the increase in HNC but does not fully explain it(29).

Overall survival estimates for HNC align with findings from the UK(20) and Europe(30,31) with cancers of the hypopharynx and nasal cavity and sinuses having lowest survival rates(32). Poorer survival rates for these cancers can be attributed to late presentation, early lymph nodal metastasis, and high cancer recurrence(33) with most patients asymptomatic until diagnosis relative to other subsites(34).

Our study revealed no consistent improvements in short-term survival and only a slight improvement in long-term survival in the UK aligning with research indicating that survival rates for oral and pharyngeal cancer have not improved in recent decades(34,35). As our study period covers up to 2021, small improvements in long term survival could be attributed to a larger reduction in smoking and alcohol and better disease management and advances in palliative care(36).

This study provides an up-to-date, analysis of HNC in the UK using two large primary care databases, spanning two decades. By presenting a comprehensive overview of long-term trends in incidence and survival across all HNC subsites, our findings extend the existing literature, much of which has focused on smaller populations or shorter time periods in the UK. Importantly, our results are in line with other studies demonstrating the value of routine primary care data for tracking evolving HNC patterns. We showed consistency across both databases supporting the robustness and generalisability of our findings. However, some variations between CPRD GOLD and Aurum, particularly for less common HNC subsites may be attributed to the databases’ underlying populations, geographic coverage(37) and differences in clinical recording practices may influence diagnostic specificity and completeness.

Our study had several limitations. We used primary care data without linkage to cancer registries, which may have introduced misclassification or delays in recording. However, linkage to cancer registries for CPRD is only available for England up to 2018 meaning the comparison with other UK regions would not be possible. Furthermore, completeness of cancer staging information has shown to be low before 2014 in linked CPRD data(38) limiting analysis of our study period if stratified by stage. However, our study is still important because staging is a function of incidence and diagnostic efforts therefore it is vital that up-to-date incidence studies are completed. However, our findings align with national cancer incidence trends, lending support to the external validity of our results and suggests that our data reflect broader population trends, despite the limitations of primary care-only records. Finally, the anatomical proximity between the different sites of HNC often leads to erroneous classification of HNC, which may result in misclassification which could change over time or by different regions. Furthermore, HNC classification used in our study does not always align with the anatomical classification used by otolaryngologists(39) meaning oral cavity and/or oropharyngeal subsites may be underreported.

## CONCLUSION

This study highlights increasing incidence of HNC in the UK are largely driven by an aging population, while specific subsites are likely influenced by additional behavioural factors. The lack of significant improvement in short-term survival and only modest gains in five-year survival emphasize the critical need for earlier diagnosis and tailored strategies to improve outcomes. Continued surveillance and research are essential to address the unique epidemiology of HNC subsites and guide prevention, diagnosis, and treatment efforts.

## Data Availability

This study is based in part on data from the Clinical Practice Research Datalink (CPRD) obtained under licence from the UK Medicines and Healthcare products Regulatory Agency. The data is provided by patients and collected by the NHS as part of their care and support. The interpretation and conclusions contained in this study are those of the author/s alone. Patient level data used in this study was obtained through an approved application-the CPRD (application number 22_001843) and is only available following an approval process-safeguard the confidentiality of patient data. Details on how to apply for data access can be found at https://cprd.com/data-access.

## ABBREVIATIONS

IR: incidence rates
HNC: head and neck cancers
KM: Kaplan-Meier
HPV: human papillomavirus

## DATA AVAILIABILITY STATEMENT

Individual patient consent was not required, as CPRD data are de-identified and provided under ethical approval from the UK Health Research Authority (HRA) and the NHS Health and Social Care Research Ethics Committee.

## CONTRIBUTIONS

Conceptualization; DN, DPA, EB

Data harmonisation and data quality assessment; AD, WYM

Formal analysis; DN

Funding acquisition; DPA

Supervision; DPA, EB, FXA-J

Interpretation of results: All authors

Roles/Writing - original draft: AMD, DN

Writing - review & editing: All authors

## REFERENCES

1. Johnson DE, Burtness B, Leemans CR, Lui VWY, Bauman JE, Grandis JR. Head and neck squamous cell carcinoma. Nature Reviews Disease Primers 2020 6:1 [Internet]. 2020 Nov 26 [cited 2025 Jul 29];6(1):1–22. Available from: https://www.nature.com/articles/s41572-020-00224-3

2. Longo DL, Chow LQM. Head and Neck Cancer. Longo DL, editor. New England Journal of Medicine [Internet]. 2020 Jan 2 [cited 2025 Jul 29];382(1):60–72. Available from: https://www.nejm.org/doi/pdf/10.1056/NEJMra1715715

3. Bray F, Laversanne M, Sung H, Ferlay J, Siegel RL, Soerjomataram I, et al. Global cancer statistics 2022: GLOBOCAN estimates of incidence and mortality worldwide for 36 cancers in 185 countries. CA Cancer J Clin [Internet]. 2024 May 1 [cited 2025 Jul 29];74(3):229–63. Available from: /doi/pdf/10.3322/caac.21834

4. Thomas SJ, Penfold CM, Waylen A, Ness AR. The changing aetiology of head and neck squamous cell cancer: A tale of three cancers? Clinical Otolaryngology [Internet]. 2018 Aug 1 [cited 2025 Jul 29];43(4):999–1003. Available from: https://pubmed.ncbi.nlm.nih.gov/29770611/

5. McCarthy CE, Field JK, Rajlawat BP, Field AE, Marcus MW. Trends and regional variation in the incidence of head and neck cancers in England: 2002 to 2011. Int J Oncol [Internet]. 2015 Jul 1 [cited 2025 Jul 29];47(1):204–10. Available from: https://pubmed.ncbi.nlm.nih.gov/25955390/

6. Mehanna H, Beech T, Nicholson T, El-Hariry I, McConkey C, Paleri V, et al. Prevalence of human papillomavirus in oropharyngeal and nonoropharyngeal head and neck cancer - Systematic review and meta-analysis of trends by time and region. Head Neck [Internet]. 2013 May [cited 2025 Jul 29];35(5):747–55. Available from: https://pubmed.ncbi.nlm.nih.gov/22267298/

7. Herrett E, Gallagher AM, Bhaskaran K, Forbes H, Mathur R, Staa T van, et al. Data Resource Profile: Clinical Practice Research Datalink (CPRD). Int J Epidemiol [Internet]. 2015 Jun 1 [cited 2025 Jul 29];44(3):827–36. Available from: 10.1093/ije/dyv098

8. Wolf A, Dedman D, Campbell J, Booth H, Lunn D, Chapman J, et al. Data resource profile: Clinical Practice Research Datalink (CPRD) Aurum. Int J Epidemiol [Internet]. 2019 Dec 1 [cited 2025 Jul 29];48(6):1740–1740g. Available from: 10.1093/ije/dyz034

9. Sanchez-Santos MT, Axson EL, Dedman D, Delmestri A. Data Resource Profile Update: CPRD GOLD. Int J Epidemiol [Internet]. 2025 Jun 11 [cited 2025 Jul 29];54(4). Available from: 10.1093/ije/dyaf077

10. Voss EA, Makadia R, Matcho A, Ma Q, Knoll C, Schuemie M, et al. Feasibility and utility of applications of the common data model to multiple, disparate observational health databases. Journal of the American Medical Informatics Association [Internet]. 2015 May 1 [cited 2025 Jul 29];22(3):553–64. Available from: https://pubmed.ncbi.nlm.nih.gov/25670757/

11. oxford-pharmacoepi/EHDENCancerIncidencePrevalence [Internet]. [cited 2025 Jul 29]. Available from: https://github.com/oxford-pharmacoepi/EHDENCancerIncidencePrevalence

12. Eurostat’s task force. Eurostat Methodologies and Working papers. Revision of the European Standard Population. Report of Eurostat’s task force. 2013 [cited 2025 Jul 29];(1346):128. Available from: http://europa.eu

13. Official Statistics | N. Ireland Cancer Registry [Internet]. [cited 2025 Jul 29]. Available from: https://www.qub.ac.uk/research-centres/nicr/CancerInformation/official-statistics/

14. Cancer incidence: 2002 to 2021 | GOV.WALES [Internet]. [cited 2025 Jul 29]. Available from: https://www.gov.wales/cancer-incidence-2002-2021

15. Cancer incidence in Scotland - to December 2022 - Cancer incidence in Scotland – Publications - Public Health Scotland [Internet]. [cited 2025 Jul 29]. Available from: https://publichealthscotland.scot/publications/cancer-incidence-in-scotland/cancer-incidence-in-scotland-to-december-2022/

16. Cancer data work programme - NDRS [Internet]. [cited 2025 Jul 29]. Available from: https://digital.nhs.uk/ndrs/our-work/ncras-work-programme

17. Raventós B, Català M, Du M, Guo Y, Black A, Inberg G, et al. IncidencePrevalence: An R package to calculate population-level incidence rates and prevalence using the OMOP common data model. Pharmacoepidemiol Drug Saf [Internet]. 2024 Jan 1 [cited 2025 Jul 29];33(1). Available from: https://pubmed.ncbi.nlm.nih.gov/37876360/

18. Therneau TM. Survival Analysis [R package survival version 3.2-13]. CRAN: Contributed Packages [Internet]. 2024 Dec 17 [cited 2025 Jul 29]; Available from: https://CRAN.R-project.org/package=survival

19. Pulte D, Brenner H. Changes in Survival in Head and Neck Cancers in the Late 20th and Early 21st Century: A Period Analysis. Oncologist [Internet]. 2010 Sep 1 [cited 2025 Jul 29];15(9):994–1001. Available from: https://pubmed.ncbi.nlm.nih.gov/20798198/

20. Gormley M, Creaney G, Schache A, Ingarfield K, Conway DI. Reviewing the epidemiology of head and neck cancer: definitions, trends and risk factors. Br Dent J [Internet]. 2022 Nov 11 [cited 2025 Jul 29];233(9):780–6. Available from: https://www.nature.com/articles/s41415-022-5166-x

21. Smith CDL, McMahon AD, Purkayastha M, Creaney G, Clements K, Inman GJ, et al. Head and neck cancer incidence is rising but the sociodemographic profile is unchanging: a population epidemiological study (2001-2020). BJC reports [Internet]. 2024 Sep 17 [cited 2025 Jul 29];2(1). Available from: https://pubmed.ncbi.nlm.nih.gov/39301277/

22. Bagnardi V, Rota M, Botteri E, Tramacere I, Islami F, Fedirko V, et al. Alcohol consumption and site-specific cancer risk: A comprehensive dose-response meta-analysis. Br J Cancer [Internet]. 2015 Feb 3 [cited 2025 Jul 29];112(3):580–93. Available from: https://pubmed.ncbi.nlm.nih.gov/25422909/

23. Mehanna H, Paleri V, West CML, Nutting C. Head and neck cancer - Part 1: Epidemiology, presentation, and prevention. BMJ (Online) [Internet]. 2010 Sep 25 [cited 2025 Jul 29];341(7774):663–6. Available from: https://pubmed.ncbi.nlm.nih.gov/20855405/

24. Hardie I, Sasso A, Holmes J, Meier PS. Understanding changes in the locations of drinking occasions in Great Britain: An age-period-cohort analysis of repeat cross-sectional market research data, 2001–2019. Drug Alcohol Rev [Internet]. 2023 Jan 1 [cited 2025 Jul 29];42(1):105–18. Available from: https://pubmed.ncbi.nlm.nih.gov/36222548/

25. Reitsma MB, Kendrick PJ, Ababneh E, Abbafati C, Abbasi-Kangevari M, Abdoli A, et al. Spatial, temporal, and demographic patterns in prevalence of smoking tobacco use and attributable disease burden in 204 countries and territories, 1990â€”2019: a systematic analysis from the Global Burden of Disease Study 2019. The Lancet [Internet]. 2021 Jun 19 [cited 2025 Jul 29];397(10292):2337–60. Available from: https://www.thelancet.com/action/showFullText?pii=S0140673621011697

26. Bruni L, Albero G, Rowley J, Alemany L, Arbyn M, Giuliano AR, et al. Global and regional estimates of genital human papillomavirus prevalence among men: a systematic review and meta-analysis. Lancet Glob Health [Internet]. 2023 Sep 1 [cited 2025 Jul 29];11(9):e1345–62. Available from: https://pubmed.ncbi.nlm.nih.gov/37591583/

27. Menezes F dos S, Fernandes GA, Antunes JLF, Villa LL, Toporcov TN. Global incidence trends in head and neck cancer for HPV-related and -unrelated subsites: A systematic review of population-based studies. Oral Oncol [Internet]. 2021 Apr 1 [cited 2025 Jul 29];115. Available from: https://pubmed.ncbi.nlm.nih.gov/33561611/

28. Kim S II, Lee JW, Eun YG, Lee YC. A SEER-based analysis of trends in HPV-associated oropharyngeal squamous cell carcinoma. Infect Agent Cancer [Internet]. 2024 Dec 1 [cited 2025 Jul 29];19(1):1–10. Available from: https://infectagentscancer.biomedcentral.com/articles/10.1186/s13027-024-00592-5

29. Schache AG, Powell NG, Cuschieri KS, Robinson M, Leary S, Mehanna H, et al. HPV-related oropharynx cancer in the United Kingdom: An evolution in the understanding of disease etiology. Cancer Res [Internet]. 2016 Nov 15 [cited 2025 Jul 29];76(22):6598–606. Available from: https://pubmed.ncbi.nlm.nih.gov/27569214/

30. Denissoff A, Huusko T, Ventelä S, Niemelä S, Routila J. Exposure to alcohol and overall survival in head and neck cancer: A regional cohort study. Head Neck [Internet]. 2022 Oct 1 [cited 2025 Jul 29];44(10):2109–17. Available from: /doi/pdf/10.1002/hed.27125

31. Cadoni G, Giraldi L, Petrelli L, Pandolfini M, Giuliani M, Paludetti G, et al. Prognostic factors in head and neck cancer: a 10-year retrospective analysis in a single-institution in Italy. Acta Otorhinolaryngologica Italica [Internet]. 2017 Dec 1 [cited 2025 Jul 29];37(6):458. Available from: https://pmc.ncbi.nlm.nih.gov/articles/PMC5782422/

32. Du E, Mazul AL, Farquhar D, Brennan P, Anantharaman D, Abedi-Ardekani B, et al. Long-term Survival in Head and Neck Cancer: Impact of Site, Stage, Smoking, and Human Papillomavirus Status. Laryngoscope [Internet]. 2019 Nov 1 [cited 2025 Jul 29];129(11):2506–13. Available from: https://pubmed.ncbi.nlm.nih.gov/30637762/

33. Hall SF, Groome PA, Irish J, O’Sullivan B. The natural history of patients with squamous cell carcinoma of the hypopharynx. Laryngoscope [Internet]. 2008 Aug [cited 2025 Jul 29];118(8):1362–71. Available from: https://pubmed.ncbi.nlm.nih.gov/18496152/

34. Sanghvi S, Khan MN, Patel NR, Yeldandi S, Baredes S, Eloy JA. Epidemiology of sinonasal squamous cell carcinoma: A comprehensive analysis of 4994 patients. Laryngoscope [Internet]. 2014 Jan 1 [cited 2025 Jul 29];124(1):76–83. Available from: /doi/pdf/10.1002/lary.24264

35. Warnakulasuriya S. Global epidemiology of oral and oropharyngeal cancer. Oral Oncol [Internet]. 2009 Apr [cited 2025 Jul 29];45(4–5):309–16. Available from: https://pubmed.ncbi.nlm.nih.gov/18804401/

36. Ganiy Opeyemi Abdulrahman J. The effect of multidisciplinary team care on cancer management. Pan Afr Med J [Internet]. 2011 Oct 26 [cited 2025 Jul 29];9(1):20. Available from: https://pmc.ncbi.nlm.nih.gov/articles/PMC3215542/

37. Mayor S. Survival rates with head and neck cancers vary widely, audit finds. BMJ [Internet]. 2015 [cited 2025 Jul 29];351:h4758. Available from: https://pubmed.ncbi.nlm.nih.gov/26340919/

38. Strongman H, Williams R, Bhaskaran K. What are the implications of using individual and combined sources of routinely collected data to identify and characterise incident site-specific cancers? a concordance and validation study using linked English electronic health records data. BMJ Open [Internet]. 2020 Aug 1 [cited 2025 Jul 29];10(8):e037719. Available from: https://bmjopen.bmj.com/content/10/8/e037719

39. Burgun A, Bodenreider O, Mougin F. Classifying diseases with respect to anatomy: a study in SNOMED CT. AMIA Annual Symposium Proceedings [Internet]. 2005 [cited 2025 Jul 29];2005:91. Available from: https://pmc.ncbi.nlm.nih.gov/articles/PMC1560776/

