## Supplement for "Incidence and Survival of Head and Neck Cancers in the United Kingdom 2000-2021"

**Supplementary Information**

### **Table 1: Clinical codelists for head and neck cancer and subsites**

The clinical codelists used for HNC and subsites is listed in the table below with the corresponding SNOMED concept ID, OMOP concept ID and concept description. Only diagnosis records alone were used to identify HNC and subtypes outcomes for this study. For the generic HNC phenotype all codelists for all subtypes were used however there were some codelists that could not be grouped into subtypes therefore were only included in the generic HNC phenotype. For example, malignant neoplasm of lip, oral cavity and pharynx cannot be placed into one subsite therefore this codelist was only used in the generic HNC codelists. We developed concept definitions using ATLAS, the OHDSI open-source platform (<https://github.com/OHDSI/atlas>). Clinical adjudicators reviewed the cohort definitions and associated concept sets.  Full details of phenotypes can be found <https://dpa-pde-oxford.shinyapps.io/EHDENCancerIncPrevCohortDiagShiny/.>

| Concept Id | Description | Concept SNOMED Code | Group |
| --- | --- | --- | --- |
| 28083 | Primary malignant neoplasm of pharynx | 93961004 | Head & Neck |
| 436045 | Malignant neoplasm of lip, oral cavity and pharynx | 271323007 | Head & Neck |
| 444224 | Overlapping malignant neoplasm of lip, oral cavity and pharynx | 109833003 | Head & Neck |
| 4095442 | Malignant neoplasm of nasal cavities, middle ear and accessory sinuses | 187828007 | Head & Neck |
| 4111021 | Overlapping malignant neoplasm of oral cavity and lips and salivary glands | 255070009 | Head & Neck |
| 4113107 | Carcinoma of lip, oral cavity and pharynx | 255069008 | Head & Neck |
| 4177243 | Malignant tumor of pharynx | 363507003 | Head & Neck |
| 4246037 | Primary malignant neoplasm of eustachian tube | 93788000 | Head & Neck |
| 4256777 | Squamous cell carcinoma of pharynx | 408649007 | Head & Neck |
| 36716486 | Primary rhabdomyosarcoma of pharynx | 722510006 | Head & Neck |
| 37117762 | Primary squamous cell carcinoma of pharynx | 733345003 | Head & Neck |
| 40486208 | Carcinoma of pharynx | 449254004 | Head & Neck |
| 434577 | Primary malignant neoplasm of hypopharyngeal aspect of aryepiglottic fold | 93829002 | hypopharynx |
| 435474 | Primary malignant neoplasm of posterior hypopharyngeal wall | 93968005 | hypopharynx |
| 436643 | Primary malignant neoplasm of anterior aspect of epiglottis | 93670003 | hypopharynx |
| 436922 | Primary malignant neoplasm of postcricoid region | 93967000 | hypopharynx |
| 438360 | Primary malignant neoplasm of vallecula | 94132005 | hypopharynx |
| 439746 | Primary malignant neoplasm of hypopharynx | 93831006 | hypopharynx |
| 440044 | Overlapping malignant neoplasm of hypopharynx | 109368005 | hypopharynx |
| 4089652 | Malignant tumor aryepiglottic fold - hypopharyngeal aspect | 187708004 | hypopharynx |
| 4091931 | Malignant neoplasm of anterior epiglottis | 187681002 | hypopharynx |
| 4092057 | Malignant tumor of pharyngeal recess | 187697007 | hypopharynx |
| 4116339 | Malignant tumor of posterior wall of hypopharynx | 303012000 | hypopharynx |
| 4177102 | Malignant tumor of vallecula | 363395000 | hypopharynx |
| 4177104 | Malignant tumor of postcricoid region | 363400004 | hypopharynx |
| 4180789 | Malignant tumor of pyriform fossa | 363401000 | hypopharynx |
| 4181342 | Malignant tumor of hypopharynx | 363399006 | hypopharynx |
| 4311479 | Primary malignant neoplasm of hypopharyngeal aspect of interarytenoid fold | 93830007 | hypopharynx |
| 40490990 | Malignant epithelial neoplasm of hypopharynx | 448665005 | hypopharynx |
| 45768871 | Primary adenocarcinoma of hypopharynx | 707395000 | hypopharynx |
| 45768882 | Primary mucoepidermoid carcinoma of hypopharynx | 707406005 | hypopharynx |
| 45768942 | Primary papillary squamous cell carcinoma of hypopharynx | 707482009 | hypopharynx |
| 45768943 | Primary undifferentiated carcinoma of hypopharynx | 707483004 | hypopharynx |
| 45768944 | Primary adenoid squamous cell carcinoma of hypopharynx | 707484005 | hypopharynx |
| 45768945 | Primary adenosquamous carcinoma of hypopharynx | 707485006 | hypopharynx |
| 45768946 | Primary basaloid carcinoma of hypopharynx | 707486007 | hypopharynx |
| 45768947 | Primary giant cell carcinoma of hypopharynx | 707487003 | hypopharynx |
| 45768948 | Primary spindle cell squamous cell carcinoma of hypopharynx | 707489000 | hypopharynx |
| 45768949 | Primary verrucous carcinoma of hypopharynx | 707490009 | hypopharynx |
| 45768951 | Primary squamous cell carcinoma of hypopharynx | 707492001 | hypopharynx |
| 45768984 | Primary squamous cell carcinoma of anterior surface of epiglottis | 707537007 | hypopharynx |
| 45768985 | Primary adenoid cystic carcinoma of hypopharynx | 707539005 | hypopharynx |
| 45769058 | Primary salivary gland type carcinoma of hypopharynx | 707627009 | hypopharynx |
| 45769059 | Overlapping squamous cell carcinoma of hypopharynx | 707628004 | hypopharynx |
| 45769108 | Primary squamous cell carcinoma of posterior wall of hypopharynx | 707686002 | hypopharynx |
| 45769121 | Primary squamous cell carcinoma of postcricoid region | 707703001 | hypopharynx |
| 45771033 | Primary squamous cell carcinoma of hypopharyngeal aspect of aryepiglottic fold | 707697002 | hypopharynx |
| 45772941 | Primary basaloid squamous cell carcinoma of hypopharynx | 707481002 | hypopharynx |
| 45773564 | Primary squamous cell carcinoma of vallecula | 707538002 | hypopharynx |
| 22839 | Overlapping malignant neoplasm of larynx | 109369002 | larynx |
| 26052 | Primary malignant neoplasm of larynx | 371995001 | larynx |
| 259755 | Primary malignant neoplasm of subglottis | 94075002 | larynx |
| 260336 | Primary malignant neoplasm of glottis | 93816002 | larynx |
| 261514 | Primary malignant neoplasm of supraglottis | 94080006 | larynx |
| 436352 | Primary malignant neoplasm of laryngeal cartilage | 109370001 | larynx |
| 4089646 | Malignant neoplasm of junctional region of epiglottis | 187685006 | larynx |
| 4091932 | Malignant neoplasm of epiglottis, free border | 187682009 | larynx |
| 4092211 | Malignant tumor of glottis | 187841006 | larynx |
| 4092212 | Malignant tumor of supraglottis | 187842004 | larynx |
| 4092213 | Malignant neoplasm of arytenoid cartilage | 187843009 | larynx |
| 4092214 | Malignant neoplasm of cuneiform cartilage | 187845002 | larynx |
| 4095309 | Malignant neoplasm of glossoepiglottic fold | 187683004 | larynx |
| 4095446 | Malignant neoplasm of cricoid cartilage | 187844003 | larynx |
| 4095447 | Malignant neoplasm of thyroid cartilage | 187846001 | larynx |
| 4110559 | Malignant tumor of anterior commissure | 254509006 | larynx |
| 4110562 | Malignant tumor of posterior commissure | 254513004 | larynx |
| 4110565 | Malignant tumor of suprahyoid epiglottis | 254517003 | larynx |
| 4110566 | Malignant tumor of infrahyoid epiglottis | 254520006 | larynx |
| 4110567 | Malignant tumor of laryngeal ventricle | 254526000 | larynx |
| 4155169 | Primary malignant neoplasm of false vocal cord | 371988005 | larynx |
| 4155170 | Primary malignant neoplasm of laryngeal aspect of aryepiglottic fold | 371994002 | larynx |
| 4157451 | Carcinoma of glottis | 372103002 | larynx |
| 4157452 | Carcinoma of subglottis | 372104008 | larynx |
| 4162122 | Primary malignant neoplasm of vocal cord | 372030009 | larynx |
| 4162136 | Carcinoma of supraglottis | 372105009 | larynx |
| 4162999 | Carcinoma of vocal cord | 372141009 | larynx |
| 4172359 | Carcinoma of larynx | 276975007 | larynx |
| 4177111 | Malignant tumor of subglottis | 363430007 | larynx |
| 4177237 | Malignant tumor of false cord | 363488008 | larynx |
| 4178968 | Malignant tumor of larynx | 363429002 | larynx |
| 4178978 | Malignant tumor of aryepiglottic fold - laryngeal aspect | 363487003 | larynx |
| 4180910 | Malignant tumor of vocal cord | 363486007 | larynx |
| 4181349 | Malignant tumor of laryngeal cartilage | 363431006 | larynx |
| 4237016 | Squamous cell carcinoma of larynx | 405822008 | larynx |
| 4246922 | Primary malignant neoplasm of epiglottis | 93784003 | larynx |
| 4247830 | Primary malignant neoplasm of laryngeal commissure | 93857009 | larynx |
| 4247831 | Primary malignant neoplasm of laryngeal surface of epiglottis | 93858004 | larynx |
| 4252536 | Squamous cell carcinoma of epiglottis | 408648004 | larynx |
| 40490009 | Transglottic malignant neoplasm of larynx | 448509007 | larynx |
| 45768839 | Primary squamous cell carcinoma of laryngeal cartilage | 707357005 | larynx |
| 45768840 | Primary squamous cell carcinoma of larynx | 707358000 | larynx |
| 45768893 | Primary undifferentiated carcinoma of larynx | 707421004 | larynx |
| 45768894 | Primary spindle cell squamous cell carcinoma of larynx | 707422006 | larynx |
| 45768895 | Primary basaloid carcinoma of larynx | 707423001 | larynx |
| 45768896 | Primary adenoid squamous cell carcinoma of larynx | 707425008 | larynx |
| 45768897 | Primary papillary squamous cell carcinoma of larynx | 707426009 | larynx |
| 45768898 | Primary verrucous carcinoma of larynx | 707427000 | larynx |
| 45768899 | Overlapping squamous cell carcinoma of larynx | 707429002 | larynx |
| 45768900 | Overlapping squamous cell carcinoma of laryngeal cartilage | 707430007 | larynx |
| 45768940 | Primary adenocarcinoma of subglottis | 707479004 | larynx |
| 45769017 | Primary squamous cell carcinoma of supraglottis | 707575007 | larynx |
| 45769018 | Primary squamous cell carcinoma of subglottis | 707576008 | larynx |
| 45769087 | Primary giant cell carcinoma of larynx | 707660007 | larynx |
| 45769089 | Primary basaloid squamous cell carcinoma of larynx | 707662004 | larynx |
| 45769091 | Primary squamous cell carcinoma of glottis | 707664003 | larynx |
| 45771018 | Primary adenosquamous cell carcinoma of larynx | 707424007 | larynx |
| 45772928 | Primary lymphoepithelial carcinoma of larynx | 707360003 | larynx |
| 132258 | Primary malignant neoplasm of frontal sinus | 93808006 | Nasal cavity & Sinus |
| 136639 | Primary malignant neoplasm of sphenoidal sinus | 94067008 | Nasal cavity & Sinus |
| 137800 | Primary malignant neoplasm of maxillary sinus | 93889000 | Nasal cavity & Sinus |
| 140046 | Primary malignant neoplasm of ethmoidal sinus | 93787005 | Nasal cavity & Sinus |
| 253977 | Overlapping malignant neoplasm of accessory sinuses | 109366009 | Nasal cavity & Sinus |
| 259748 | Primary malignant neoplasm of accessory sinus | 93659005 | Nasal cavity & Sinus |
| 435190 | Primary malignant neoplasm of pyriform sinus | 93978008 | Nasal cavity & Sinus |
| 438367 | Primary malignant neoplasm of nasal cavity | 93917007 | Nasal cavity & Sinus |
| 4092209 | Malignant neoplasm of nasal conchae | 187830009 | Nasal cavity & Sinus |
| 4092210 | Malignant tumor of nasal vestibule | 187831008 | Nasal cavity & Sinus |
| 4110434 | Malignant tumor of inferior turbinate | 254478004 | Nasal cavity & Sinus |
| 4110435 | Malignant tumor of posterior margin of nasal septum and choanae | 254484001 | Nasal cavity & Sinus |
| 4111022 | Malignant tumor of lateral nasal wall | 255075004 | Nasal cavity & Sinus |
| 4112593 | Malignant tumor of middle turbinate | 254481009 | Nasal cavity & Sinus |
| 4177107 | Malignant tumor of nasal cavity | 363422006 | Nasal cavity & Sinus |
| 4177109 | Malignant tumor of ethmoid sinus | 363426009 | Nasal cavity & Sinus |
| 4177110 | Malignant tumor of sphenoid sinus | 363428005 | Nasal cavity & Sinus |
| 4181347 | Malignant tumor of maxillary sinus | 363425008 | Nasal cavity & Sinus |
| 4181348 | Malignant tumor of frontal sinus | 363427000 | Nasal cavity & Sinus |
| 4181486 | Malignant tumor of nasal sinuses | 363506007 | Nasal cavity & Sinus |
| 4246138 | Primary malignant neoplasm of palatine bone | 93936002 | Nasal cavity & Sinus |
| 4247843 | Primary malignant neoplasm of nasal concha | 93918002 | Nasal cavity & Sinus |
| 4309385 | Carcinoma of nasal meatus | 422758009 | Nasal cavity & Sinus |
| 4312032 | Primary malignant neoplasm of vestibule of nose | 94139001 | Nasal cavity & Sinus |
| 36715799 | Primary adenocarcinoma of nasal cavity | 721560002 | Nasal cavity & Sinus |
| 36715835 | Primary squamous cell carcinoma of overlapping lesion of accessory sinuses | 721605003 | Nasal cavity & Sinus |
| 36715836 | Primary adenocarcinoma of overlapping lesion of accessory sinuses | 721606002 | Nasal cavity & Sinus |
| 36716941 | Primary squamous cell carcinoma of nasal cavity | 723182009 | Nasal cavity & Sinus |
| 40492934 | Carcinoma of nasal cavity | 448990005 | Nasal cavity & Sinus |
| 42536528 | Primary malignant neuroepitheliomatous neoplasm of nasal cavity | 735450006 | Nasal cavity & Sinus |
| 44783808 | Undifferentiated carcinoma of nasal sinus | 697993003 | Nasal cavity & Sinus |
| 45768824 | Primary adenocarcinoma of accessory sinus | 707337006 | Nasal cavity & Sinus |
| 45768828 | Primary adenocarcinoma of ethmoidal sinus | 707342003 | Nasal cavity & Sinus |
| 45768829 | Primary adenocarcinoma of frontal sinus | 707343008 | Nasal cavity & Sinus |
| 45768830 | Primary adenocarcinoma of sphenoidal sinus | 707344002 | Nasal cavity & Sinus |
| 45768831 | Primary carcinoma of ethmoidal sinus | 707346000 | Nasal cavity & Sinus |
| 45768832 | Primary carcinoma of maxillary sinus | 707347009 | Nasal cavity & Sinus |
| 45768833 | Primary carcinoma of frontal sinus | 707349007 | Nasal cavity & Sinus |
| 45768836 | Primary squamous cell carcinoma of accessory sinus | 707353009 | Nasal cavity & Sinus |
| 45768837 | Primary squamous cell carcinoma of sphenoidal sinus | 707355002 | Nasal cavity & Sinus |
| 45768838 | Primary squamous cell carcinoma of frontal sinus | 707356001 | Nasal cavity & Sinus |
| 45769122 | Primary squamous cell carcinoma of pyriform sinus | 707704007 | Nasal cavity & Sinus |
| 45771012 | Primary adenocarcinoma of maxillary sinus | 707339009 | Nasal cavity & Sinus |
| 45771013 | Primary carcinoma of accessory sinus | 707345001 | Nasal cavity & Sinus |
| 45771015 | Primary squamous cell carcinoma of maxillary sinus | 707354003 | Nasal cavity & Sinus |
| 45772927 | Primary carcinoma of sphenoidal sinus | 707348004 | Nasal cavity & Sinus |
| 45773563 | Primary squamous cell carcinoma of ethmoidal sinus | 707359008 | Nasal cavity & Sinus |
| 432558 | Overlapping malignant neoplasm of nasopharynx | 109367000 | nasopharynx |
| 436344 | Primary malignant neoplasm of posterior wall of nasopharynx | 93970001 | nasopharynx |
| 437226 | Malignant neoplasm of nasopharyngeal wall | 240163000 | nasopharynx |
| 438080 | Primary malignant neoplasm of anterior wall of nasopharynx | 93674007 | nasopharynx |
| 438692 | Primary malignant neoplasm of lateral wall of nasopharynx | 93861003 | nasopharynx |
| 441223 | Primary malignant neoplasm of superior wall of nasopharynx | 94078000 | nasopharynx |
| 761907 | Primary adenocarcinoma of nasopharynx | 184861000119102 | nasopharynx |
| 4084147 | Metastatic malignant neoplasm to nasopharynx | 241861008 | nasopharynx |
| 4089647 | Malignant tumor of adenoid | 187694000 | nasopharynx |
| 4089649 | Malignant neoplasm of floor of nasopharynx | 187701005 | nasopharynx |
| 4094722 | Malignant tumor of posterior wall of nasopharynx | 187693006 | nasopharynx |
| 4095312 | Malignant tumor of nasopharynx | 187692001 | nasopharynx |
| 4095313 | Malignant tumor of anterior wall of nasopharynx | 187700006 | nasopharynx |
| 4116237 | Malignant tumor of nasal cavity and nasopharynx | 255074000 | nasopharynx |
| 4177103 | Malignant tumor of lateral wall of nasopharynx | 363398003 | nasopharynx |
| 4306501 | Undifferentiated carcinoma of nasopharynx | 422541001 | nasopharynx |
| 4307152 | Adenocarcinoma of nasopharynx | 423106003 | nasopharynx |
| 4309248 | Squamous cell carcinoma of nasopharynx | 422691006 | nasopharynx |
| 36684473 | Primary malignant neoplasm of nasopharynx | 226521000119108 | nasopharynx |
| 36716501 | Primary malignant epithelial neoplasm of nasopharynx | 722529000 | nasopharynx |
| 37018571 | Primary adenoid cystic carcinoma of nasopharynx | 7391000119103 | nasopharynx |
| 40486201 | Nasopharyngeal carcinoma | 449248000 | nasopharynx |
| 44782582 | Keratinizing squamous cell carcinoma of nasopharynx | 698011002 | nasopharynx |
| 44783851 | Undifferentiated nonkeratinizing carcinoma of nasopharynx | 698048006 | nasopharynx |
| 45768977 | Primary squamous cell carcinoma of nasopharynx | 707528007 | nasopharynx |
| 45769123 | Nonkeratinizing carcinoma of the nasopharynx | 707705008 | nasopharynx |
| 46271011 | Malignant neoplasm of superior wall of nasopharynx | 709031009 | nasopharynx |
| 25189 | Malignant tumor of oral cavity | 363505006 | Oral Cavity |
| 132565 | Primary malignant neoplasm of vermilion border of lower lip | 372026006 | Oral Cavity |
| 132832 | Primary malignant neoplasm of inner aspect of lip | 93835002 | Oral Cavity |
| 133710 | Overlapping malignant neoplasm of lip | 109822001 | Oral Cavity |
| 133969 | Primary malignant neoplasm of vermilion border of lip | 94135007 | Oral Cavity |
| 134290 | Primary malignant neoplasm of palate | 372002009 | Oral Cavity |
| 134579 | Primary malignant neoplasm of buccal mucosa | 371976008 | Oral Cavity |
| 135750 | Primary malignant neoplasm of floor of mouth | 93802007 | Oral Cavity |
| 137219 | Primary malignant neoplasm of inner aspect of lower lip | 93836001 | Oral Cavity |
| 138074 | Primary malignant neoplasm of vermilion border of upper lip | 372027002 | Oral Cavity |
| 138351 | Primary malignant neoplasm of inner aspect of upper lip | 93837005 | Oral Cavity |
| 140950 | Primary malignant neoplasm of gum | 371990006 | Oral Cavity |
| 140955 | Overlapping malignant neoplasm of floor of mouth | 109830000 | Oral Cavity |
| 140958 | Kaposi's sarcoma of palate | 109388009 | Oral Cavity |
| 254282 | Primary malignant neoplasm of soft palate | 94049001 | Oral Cavity |
| 255192 | Primary malignant neoplasm of commissure of lip | 371981004 | Oral Cavity |
| 261808 | Primary malignant neoplasm of vestibule of mouth | 94138009 | Oral Cavity |
| 433704 | Primary malignant neoplasm of retromolar area | 93989001 | Oral Cavity |
| 434285 | Primary malignant neoplasm of uvula | 94129007 | Oral Cavity |
| 435478 | Primary malignant neoplasm of upper gum | 372022008 | Oral Cavity |
| 438694 | Primary malignant neoplasm of hard palate | 371991005 | Oral Cavity |
| 438982 | Primary malignant neoplasm of anterior portion of floor of mouth | 93672006 | Oral Cavity |
| 439404 | Primary malignant neoplasm of oral cavity | 372001002 | Oral Cavity |
| 439738 | Primary malignant neoplasm of sublingual gland | 94076001 | Oral Cavity |
| 440335 | Primary malignant neoplasm of lower gum | 371997009 | Oral Cavity |
| 440344 | Primary malignant neoplasm of lateral portion of floor of mouth | 93860002 | Oral Cavity |
| 4001170 | Overlapping malignant neoplasm of palate | 109831001 | Oral Cavity |
| 4081190 | Squamous cell carcinoma of floor of mouth | 276954004 | Oral Cavity |
| 4089516 | Malignant neoplasm of lower lip, external | 187604008 | Oral Cavity |
| 4089520 | Malignant tumor of labial mucosa | 187622006 | Oral Cavity |
| 4089530 | Malignant tumor of lateral floor of mouth | 187653008 | Oral Cavity |
| 4089644 | Malignant neoplasm of junction of hard and soft palate | 187666008 | Oral Cavity |
| 4090216 | Malignant tumor of upper labial mucosa | 187606005 | Oral Cavity |
| 4090217 | Malignant tumor of frenum of lower lip | 187614004 | Oral Cavity |
| 4090224 | Malignant tumor of anterior floor of mouth | 187652003 | Oral Cavity |
| 4090226 | Malignant tumor of vestibule of mouth | 187658004 | Oral Cavity |
| 4090227 | Malignant tumor of lower buccal sulcus | 187660002 | Oral Cavity |
| 4090228 | Malignant tumor of upper labial sulcus | 187661003 | Oral Cavity |
| 4090229 | Malignant tumor of lower labial sulcus | 187662005 | Oral Cavity |
| 4093012 | Malignant neoplasm of upper lip, lipstick area | 187601000 | Oral Cavity |
| 4093646 | Malignant tumor of frenum of upper lip | 187608006 | Oral Cavity |
| 4093647 | Malignant neoplasm of lower lip, buccal aspect | 187613005 | Oral Cavity |
| 4094716 | Malignant tumor of upper buccal sulcus | 187659007 | Oral Cavity |
| 4094720 | Malignant tumor of tonsillar pillar | 187675005 | Oral Cavity |
| 4110417 | Carcinoma of vermilion border of lower lip | 254390001 | Oral Cavity |
| 4110422 | Carcinoma of lower gum | 254425003 | Oral Cavity |
| 4110424 | Carcinoma of lateral part of floor of mouth | 254431000 | Oral Cavity |
| 4110425 | Carcinoma of uvula | 254436005 | Oral Cavity |
| 4110428 | Carcinoma of lower buccal sulcus | 254445006 | Oral Cavity |
| 4110430 | Carcinoma of upper labial sulcus | 254450000 | Oral Cavity |
| 4111645 | Carcinoma of frenum of lip | 254393004 | Oral Cavity |
| 4111653 | Carcinoma of upper gum | 254424004 | Oral Cavity |
| 4111654 | Carcinoma of anterior part of floor of mouth | 254427006 | Oral Cavity |
| 4111775 | Carcinoma of hard palate | 254434008 | Oral Cavity |
| 4111776 | Squamous cell carcinoma of buccal mucosa | 254437001 | Oral Cavity |
| 4111777 | Carcinoma of upper buccal sulcus | 254441002 | Oral Cavity |
| 4111783 | Carcinoma of retromolar area | 254457002 | Oral Cavity |
| 4112451 | Carcinoma of vermilion border of upper lip | 254389005 | Oral Cavity |
| 4112453 | Carcinoma of frenum of upper lip | 254398008 | Oral Cavity |
| 4112455 | Carcinoma of frenum of lower lip | 254402004 | Oral Cavity |
| 4112456 | Carcinoma of commissure of lip | 254404003 | Oral Cavity |
| 4112598 | Malignant tumor of inferior surface of soft palate | 254503007 | Oral Cavity |
| 4113108 | Squamous cell carcinoma of lip | 255071008 | Oral Cavity |
| 4115130 | Carcinoma of soft palate | 254435009 | Oral Cavity |
| 4115135 | Carcinoma of lower labial sulcus | 254454009 | Oral Cavity |
| 4118988 | Malignant tumor of frenum of lip | 302815008 | Oral Cavity |
| 4145095 | Squamous cell carcinoma of mouth | 307502000 | Oral Cavity |
| 4150793 | Malignant tumor of lower labial mucosa | 271568003 | Oral Cavity |
| 4153887 | Carcinoma of lip | 269515006 | Oral Cavity |
| 4155171 | Primary malignant neoplasm of lip | 371996000 | Oral Cavity |
| 4166769 | Palate carcinoma | 274084007 | Oral Cavity |
| 4169288 | Squamous cell carcinoma of mucous membrane of lower lip | 418372008 | Oral Cavity |
| 4170451 | Malignant tumor of lipstick area of lip | 275399006 | Oral Cavity |
| 4174595 | Squamous cell carcinoma of palate | 276962007 | Oral Cavity |
| 4174910 | Squamous cell carcinoma of gum | 276953005 | Oral Cavity |
| 4177100 | Malignant tumor of commissure of lip | 363374005 | Oral Cavity |
| 4177101 | Malignant tumor of floor of mouth | 363385007 | Oral Cavity |
| 4178961 | Malignant tumor of vermilion border of lower lip | 363373004 | Oral Cavity |
| 4178963 | Malignant tumor of gum | 363382005 | Oral Cavity |
| 4178964 | Malignant tumor of palate | 363390005 | Oral Cavity |
| 4178965 | Malignant tumor of retromolar area | 363391009 | Oral Cavity |
| 4180779 | Malignant tumor of lip | 363348004 | Oral Cavity |
| 4180782 | Malignant tumor of vermilion border of upper lip | 363372009 | Oral Cavity |
| 4180786 | Malignant tumor of buccal mucosa | 363386008 | Oral Cavity |
| 4180787 | Malignant tumor of hard palate | 363387004 | Oral Cavity |
| 4181334 | Malignant tumor of upper gingiva | 363383000 | Oral Cavity |
| 4181335 | Malignant tumor of lower gingiva | 363384006 | Oral Cavity |
| 4181337 | Malignant tumor of uvula | 363389001 | Oral Cavity |
| 4225833 | Malignant tumor of vermilion border of lip | 421249001 | Oral Cavity |
| 4246015 | Primary malignant neoplasm of alveolar ridge mucosa | 93667002 | Oral Cavity |
| 4300554 | Verrucous carcinoma of oral cavity | 403889000 | Oral Cavity |
| 4302482 | Squamous cell carcinoma of mucous membrane of upper lip | 419240004 | Oral Cavity |
| 4311283 | Malignant neoplasm of gum and contiguous sites | 423691004 | Oral Cavity |
| 4311476 | Primary malignant neoplasm of gingival mucosa | 93812000 | Oral Cavity |
| 4311719 | Primary Kaposi's sarcoma of oral cavity | 424779008 | Oral Cavity |
| 35610411 | Overlapping malignant neoplasm of mouth | 1092831000000100 | Oral Cavity |
| 36715796 | Primary adenocarcinoma of palate | 721556000 | Oral Cavity |
| 36716485 | Primary rhabdomyosarcoma of oral cavity | 722509001 | Oral Cavity |
| 37018880 | Primary squamous cell carcinoma of vermilion border of lip | 242951000119108 | Oral Cavity |
| 37116583 | Primary squamous cell carcinoma of oral cavity | 733343005 | Oral Cavity |
| 37116584 | Primary squamous cell carcinoma of lip | 733344004 | Oral Cavity |
| 40486174 | Malignant neoplasm of alveolus of maxilla | 449223003 | Oral Cavity |
| 40486214 | Malignant neoplasm of alveolus dentalis | 449260004 | Oral Cavity |
| 40487580 | Malignant epithelial neoplasm of alveolus dentalis | 449472007 | Oral Cavity |
| 40488048 | Malignant neoplasm of alveolus of mandible | 449578008 | Oral Cavity |
| 40492985 | Malignant neoplasm of anterior and lateral floor of mouth | 449034009 | Oral Cavity |
| 40493495 | Carcinoma of floor of mouth | 449156009 | Oral Cavity |
| 31509 | Primary malignant neoplasm of tonsil | 372020000 | oropharynx |
| 432833 | Primary malignant neoplasm of oropharynx | 93933005 | oropharynx |
| 433709 | Primary malignant neoplasm of tonsillar fossa | 94102002 | oropharynx |
| 438691 | Overlapping malignant neoplasm of oropharynx | 109832008 | oropharynx |
| 439739 | Primary malignant neoplasm of posterior wall of oropharynx | 93971002 | oropharynx |
| 440047 | Primary malignant neoplasm of tonsillar pillar | 94103007 | oropharynx |
| 440345 | Primary malignant neoplasm of lateral wall of oropharynx | 93862005 | oropharynx |
| 4002498 | Overlapping malignant neoplasm of tonsil | 110013004 | oropharynx |
| 4094721 | Malignant tumor of posterior wall of oropharynx | 187688008 | oropharynx |
| 4094850 | Malignant neoplasm of posterior pharynx | 187709007 | oropharynx |
| 4115137 | Malignant tumor of anterior pillar of fauces | 254459004 | oropharynx |
| 4158909 | Tonsil carcinoma | 274085008 | oropharynx |
| 4180788 | Malignant tumor of tonsillar fossa | 363394001 | oropharynx |
| 4181336 | Malignant tumor of soft palate | 363388009 | oropharynx |
| 4181338 | Malignant tumor of oropharynx | 363392002 | oropharynx |
| 4181339 | Malignant tumor of tonsil | 363393007 | oropharynx |
| 4246793 | Primary malignant neoplasm of adenoid | 93662008 | oropharynx |
| 4308627 | Adenoid cystic carcinoma of oropharynx | 423318000 | oropharynx |
| 4309545 | Squamous cell carcinoma of oropharynx | 423464009 | oropharynx |
| 36715837 | Primary undifferentiated carcinoma of oropharynx | 721607006 | oropharynx |
| 36716502 | Primary squamous cell carcinoma of pharyngeal tonsil | 722530005 | oropharynx |
| 37018947 | Primary squamous cell carcinoma of palatine tonsil | 18121000119104 | oropharynx |
| 40488898 | Malignant epithelial neoplasm of oropharynx | 448214005 | oropharynx |
| 40492021 | Malignant neoplasm of lateral wall of oropharynx | 448868009 | oropharynx |
| 45768872 | Primary oxyphilic adenocarcinoma of oropharynx | 707396004 | oropharynx |
| 45768873 | Primary basal cell adenocarcinoma of oropharynx | 707397008 | oropharynx |
| 45768874 | Primary polymorphous low grade adenocarcinoma of oropharynx | 707398003 | oropharynx |
| 45768875 | Primary papillary adenocarcinoma of oropharynx | 707399006 | oropharynx |
| 45768876 | Primary mucinous adenocarcinoma of oropharynx | 707400004 | oropharynx |
| 45768877 | Primary clear cell adenocarcinoma of oropharynx | 707401000 | oropharynx |
| 45768878 | Primary adenocarcinoma of oropharynx | 707402007 | oropharynx |
| 45768978 | Overlapping squamous cell carcinoma of oropharynx | 707529004 | oropharynx |
| 45768980 | Primary squamous cell carcinoma of posterior wall of oropharynx | 707532001 | oropharynx |
| 45768982 | Primary squamous cell carcinoma of lateral wall of oropharynx | 707535004 | oropharynx |
| 45769021 | Primary basaloid carcinoma of oropharynx | 707580003 | oropharynx |
| 45769022 | Primary papillary squamous cell carcinoma of oropharynx | 707581004 | oropharynx |
| 45769023 | Primary spindle cell squamous cell carcinoma of oropharynx | 707582006 | oropharynx |
| 45769024 | Primary adenosquamous carcinoma of oropharynx | 707583001 | oropharynx |
| 45769026 | Primary squamous cell carcinoma of oropharynx | 707585008 | oropharynx |
| 45769027 | Primary myoepithelial carcinoma of oropharynx | 707586009 | oropharynx |
| 45769028 | Primary carcinoma ex pleomorphic adenoma of oropharynx | 707587000 | oropharynx |
| 45769029 | Primary epithelial-myoepithelial carcinoma of oropharynx | 707588005 | oropharynx |
| 45769030 | Primary acinar cell carcinoma of oropharynx | 707590006 | oropharynx |
| 45769031 | Primary mucoepidermoid carcinoma of oropharynx | 707591005 | oropharynx |
| 45769032 | Primary infiltrating duct carcinoma of oropharynx | 707592003 | oropharynx |
| 45771025 | Primary cystadenocarcinoma of oropharynx | 707589002 | oropharynx |
| 45771026 | Primary salivary gland-type tumor of oropharynx | 707593008 | oropharynx |
| 45772951 | Primary basaloid squamous cell carcinoma of oropharynx | 707579001 | oropharynx |
| 22557 | Malignant tumor of submandibular gland | 363380002 | Salivary Gland |
| 28356 | Overlapping malignant neoplasm of major salivary gland | 109824000 | Salivary Gland |
| 434588 | Primary malignant neoplasm of parotid gland | 372004005 | Salivary Gland |
| 439745 | Primary malignant neoplasm of salivary gland duct | 109828002 | Salivary Gland |
| 4110432 | Carcinoma of submandibular gland | 254465004 | Salivary Gland |
| 4112585 | Carcinoma of sublingual gland | 254466003 | Salivary Gland |
| 4115138 | Carcinoma of parotid gland | 254462001 | Salivary Gland |
| 4116235 | Malignant tumor of salivary gland | 255072001 | Salivary Gland |
| 4162861 | Primary malignant neoplasm of minor salivary gland | 372000001 | Salivary Gland |
| 4169627 | Squamous cell carcinoma of oral mucous membrane | 419842002 | Salivary Gland |
| 4180784 | Malignant tumor of parotid gland | 363379000 | Salivary Gland |
| 4180785 | Malignant tumor of sublingual gland | 363381003 | Salivary Gland |
| 4180909 | Malignant tumor of minor salivary gland | 363485006 | Salivary Gland |
| 4181333 | Malignant tumor of major salivary gland | 363378008 | Salivary Gland |
| 4247836 | Primary malignant neoplasm of major salivary gland | 93883004 | Salivary Gland |
| 4307266 | Adenoid cystic carcinoma of submandibular gland | 423189008 | Salivary Gland |
| 4308149 | Mucoepidermoid carcinoma of parotid gland | 423793008 | Salivary Gland |
| 4309400 | Adenoid cystic carcinoma of salivary gland | 422833009 | Salivary Gland |
| 4310565 | Mucoepidermoid carcinoma of submandibular gland | 423424005 | Salivary Gland |
| 4311287 | Mucoepidermoid carcinoma of salivary gland | 423708008 | Salivary Gland |
| 4312691 | Primary malignant neoplasm of submaxillary gland | 94077005 | Salivary Gland |
| 4312798 | Carcinoma ex pleomorphic adenoma of parotid gland | 425127006 | Salivary Gland |
| 4312929 | Polymorphous low grade adenocarcinoma of salivary gland | 423038006 | Salivary Gland |
| 4313754 | Adenoid cystic carcinoma of parotid gland | 423615009 | Salivary Gland |
| 4314314 | Malignant mixed tumor of salivary gland | 425225007 | Salivary Gland |
| 36715797 | Primary adenocarcinoma of parotid gland | 721557009 | Salivary Gland |
| 36717226 | Primary squamous cell carcinoma of parotid gland | 722674001 | Salivary Gland |
| 37204489 | Malignant epithelial neoplasm of salivary gland | 783155007 | Salivary Gland |
| 42537751 | Primary adenocarcinoma of sublingual gland | 737308008 | Salivary Gland |
| 42537752 | Primary adenocarcinoma of submandibular gland | 737309000 | Salivary Gland |
| 42537753 | Primary squamous cell carcinoma of submandibular gland | 737310005 | Salivary Gland |
| 42539700 | Primary squamous cell carcinoma of sublingual gland | 737311009 | Salivary Gland |
| 256633 | Primary malignant neoplasm of base of tongue | 93687001 | Tongue |
| 434289 | Primary malignant neoplasm of lingual tonsil | 93868009 | Tongue |
| 434587 | Primary malignant neoplasm of dorsal surface of tongue | 93773005 | Tongue |
| 436042 | Primary malignant neoplasm of anterior two-thirds of tongue | 371968006 | Tongue |
| 436043 | Overlapping malignant neoplasm of tongue | 109823006 | Tongue |
| 437220 | Primary malignant neoplasm of ventral surface of tongue | 94134006 | Tongue |
| 437498 | Primary malignant neoplasm of tongue | 94101009 | Tongue |
| 440036 | Primary malignant neoplasm of junctional zone of tongue | 93848003 | Tongue |
| 440655 | Malignant neoplasm of tongue, tip and lateral border | 187637005 | Tongue |
| 4089524 | Malignant neoplasm of base of tongue dorsal surface | 187631006 | Tongue |
| 4089526 | Malignant tumor of junctional zone of tongue | 187644001 | Tongue |
| 4093141 | Malignant tumor of anterior two-thirds of tongue - ventral surface | 187640005 | Tongue |
| 4093649 | Malignant tumor of anterior two-thirds of tongue - dorsal surface | 187634003 | Tongue |
| 4093650 | Malignant neoplasm of midline of tongue | 187635002 | Tongue |
| 4093651 | Malignant tumor of frenum linguae | 187641009 | Tongue |
| 4110420 | Malignant tumor of anterior two-thirds of tongue - lateral margin | 254408000 | Tongue |
| 4111649 | Malignant tumor of tip of tongue | 254412006 | Tongue |
| 4112580 | Carcinoma of frenum linguae | 254417000 | Tongue |
| 4112581 | Carcinoma of lingual tonsil | 254423005 | Tongue |
| 4149844 | Tongue carcinoma | 269516007 | Tongue |
| 4156114 | Primary malignant neoplasm of border of tongue | 371975007 | Tongue |
| 4158473 | Carcinoma of base of tongue | 271943005 | Tongue |
| 4162118 | Malignant neoplasm of border of tongue | 371974006 | Tongue |
| 4168069 | Carcinoma of tongue base - dorsal surface | 275490009 | Tongue |
| 4170450 | Carcinoma of anterior two-thirds of tongue - dorsal surface | 275396004 | Tongue |
| 4173799 | Carcinoma ventral surface of tongue | 275394001 | Tongue |
| 4173800 | Carcinoma anterior 2/3 tongue ventrum | 275395000 | Tongue |
| 4173801 | Carcinoma of midline of tongue | 275397008 | Tongue |
| 4174593 | Squamous cell carcinoma of tongue | 276952000 | Tongue |
| 4177098 | Malignant tumor of anterior two-thirds of tongue | 363360003 | Tongue |
| 4178962 | Malignant tumor of tongue | 363375006 | Tongue |
| 4180783 | Malignant tumor of lingual tonsil | 363377003 | Tongue |
| 4181332 | Malignant tumor of base of tongue | 363376007 | Tongue |
| 4310135 | Primary sarcoma of tongue | 424849005 | Tongue |
| 35610172 | Malignant neoplasm of ventral surface of tongue | 1090271000000100 | Tongue |
| 36716610 | Primary squamous cell carcinoma of base of tongue | 722672002 | Tongue |
| 36717352 | Primary squamous cell carcinoma of lingual tonsil | 722673007 | Tongue |
| 40381321 | Malignant neoplasm of dorsal surface of tongue | 187633009 | Tongue |

### **Table 2: Population attrition showing eligible patients for study from each database for HNC**

| **N** | **Reason** | **N excluded** | **Database** |
| --- | --- | --- | --- |
| 39999011 | Starting population |  | Aurum |
| 39999011 | Missing year of birth | 0 |  |
| 39999011 | Missing sex | 0 |  |
| 34833388 | Cannot satisfy age criteria during the study period based on year of birth | 5165623 |  |
| 29190480 | No observation time available during study period | 5642908 |  |
| 29190480 | Doesn't satisfy age criteria during the study period | 0 |  |
| 25483313 | Prior history requirement not fulfilled during study period | 3707167 |  |
| 24340860 | No observation time available after applying age and prior history criteria | 1142453 |  |
| 24340860 | Starting analysis population |  |  |
| 24340860 | Estimating prevalence |  |  |
| 24335457 | Excluded due to prior event (do not pass outcome washout during study period) | 5403 |  |
| 24335457 | Estimating incidence |  |  |
| 21356 | With a cancer diagnosis | 24314101 |  |
| 21255 | Cancer diagnosis not on same date as death | 101 |  |
| 21255 | Estimating survival |  |  |
| 17054819 | Starting population |  | GOLD |
| 17054819 | Missing year of birth | 0 |  |
| 17054819 | Missing sex | 0 |  |
| 15210165 | Cannot satisfy age criteria during the study period based on year of birth | 1844654 |  |
| 13978229 | No observation time available during study period | 1231936 |  |
| 13978229 | Doesn't satisfy age criteria during the study period | 0 |  |
| 12254874 | Prior history requirement not fulfilled during study period | 1723355 |  |
| 11388117 | No observation time available after applying age and prior history criteria | 866757 |  |
| 11388117 | Starting analysis population |  |  |
| 11388117 | Estimating prevalence |  |  |
| 11386416 | Excluded due to prior event (do not pass outcome washout during study period) | 1701 |  |
| 11386416 | Estimating incidence |  |  |
| 12455 | With a cancer diagnosis | 11373927 |  |
| 12381 | Cancer diagnosis not on same date as death | 74 |  |
| 12489 | Estimating survival |  |  |

### **Table 3 Baseline characteristics of HNC patients at the time of diagnosis for CPRD Aurum.**

| **Database** | **Aurum** |
| --- | --- |
| **Number of patients** | 21356 |
| **Sex: Male (N[%])** | 14932 (69.9%) |
| **Age (Median [IQR])** | 64 (55 to 73) |
| **Age Groups N (%)** |  |
| 18-29 | 129 (0.6%) |
| 30-39 | 470 (2.2%) |
| 40-49 | 1957 (9.2%) |
| 50-59 | 5236 (24.5%) |
| 60-69 | 6297 (29.5%) |
| 70-79 | 4630 (21.7%) |
| 80-89 | 2231 (10.4%) |
| 90+ | 406 (1.90%) |
| **Prior history, days** |  |
| median [p25 - p75] | 5717.5 (2,657 to 9,465) |
| **General conditions (any time prior)** |  |
| Cerebrovascular disease | 1246 (5.8%) |
| Chronic liver disease | 343 (1.6%) |
| Chronic obstructive lung disease | 2199 (10.3%) |
| Coronary arteriosclerosis | 209 (1.0%) |
| Diabetes mellitus | 2271 (10.6%) |
| Gastroesophageal reflux disease | 626 (2.9%) |
| Gastrointestinal haemorrhage | 1431 (6.7%) |
| Heart disease | 3710 (17.4%) |
| Heart failure | 520 (2.4%) |
| Hepatitis C | 56 (0.3%) |
| Hyperlipidemia | 1922 (9.0%) |
| Hypertensive disorder | 7041 (33.0%) |
| Ischemic heart disease | 2190 (10.3%) |
| Lesion of liver | 636 (3.0%) |
| Obesity | 573 (2.7%) |
| Osteoarthritis | 4320 (20.2%) |
| Peripheral vascular disease | 659 (3.1%) |
| Renal impairment | 1790 (8.4%) |
| Venous thrombosis | 795 (3.70%) |
| Visual system disorder | 7008 (32.8%) |

### **Table 4 Baseline characteristics per HNC subsites for CPRD GOLD**

| **Database** | **Hypopharynx** | **Larynx** | **Nasal Cavity & Sinus** | **Nasopharynx** | **Oral Cavity** | **Oropharynx** | **Salivary Gland** | **Tongue** |
| --- | --- | --- | --- | --- | --- | --- | --- | --- |
| **Number of patients** | 818 | 2797 | 258 | 372 | 1973 | 2321 | 761 | 2499 |
| **Sex: Male (N[%])** | 67 (59 to 74) | 67 (59 to 75) | 67 (55 to 74) | 60 (52 to 71) | 65 (56 to 75) | 61 (54 to 68) | 69 (55 to 79) | 62 (55 to 71) |
| **Age (Years) (Median [IQR])** | 627 (76.7%) | 2263 (80.9%) | 160 (62.0%) | 253 (68.0%) | 1140 (57.8%) | 1721 (74.1%) | 451 (59.3%) | 1650 (66.0%) |
| **Age Groups N (%)** | | | | | | | | |
| **18-29** | 0 | 13 (0.5%) | <5 | 6 (1.6%) | 28 (1.4%) | <5 | 22 (2.9%) | 14 (0.6%) |
| **30-39** | <5 | 27 (1.0%) | 11 (4.3%) | 19 (5.1%) | 62 (3.1%) | 18 (0.8%) | 37 (4.9%) | 54 (2.2%) |
| **40-49** | 38 (4.6%) | 134 (4.8%) | 24 (9.3%) | 39 (10.5%) | 175 (8.9%) | 263 (11.3%) | 73 (9.6%) | 252 (10.1%) |
| **50-59** | 171 (20.9%) | 563 (20.1%) | 49 (19.0%) | 119 (32.0%) | 417 (21.1%) | 753 (32.4%) | 116 (15.2%) | 672 (26.9%) |
| **60-69** | 277 (33.9%) | 909 (32.5%) | 63 (24.4%) | 80 (21.5%) | 539 (27.3%) | 766 (33.0%) | 151 (19.8%) | 772 (30.9%) |
| **70-79** | 233 (28.5%) | 808 (28.9%) | 79 (30.6%) | 68 (18.3%) | 435 (22.0%) | 387 (16.7%) | 182 (23.9%) | 478 (19.1%) |
| **80-89** | 89 (10.9%) | 304 (10.9%) | 30 (11.6%) | 34 (9.1%) | 264 (13.4%) | 114 (4.9%) | 139 (18.3%) | 219 (8.8%) |
| **90+** | 9 (1.1%) | 39 (1.4%) | <5 | 7 (1.9%) | 53 (2.7%) | 16 (0.7%) | 41 (5.4%) | 38 (1.5%) |
| **Prior history, days** | | | | | | | | |
| **median [IQR]** | 3496 (1,916 to 5,214) | 3350 (1,805 to 5,071) | 3491 (1,931 to 5,407) | 3365.5 (1,883 to 5,174) | 3318 (1,786 to 5,012) | 3597 (2,075 to 5,354) | 3466 (1,905 to 5,354) | 3410 (1,847 to 5,170) |
| **Smoking Status (5 years prior)** | | | | | | | | |
| **Non-Smoker** | 149 (18.2%) | 441 (15.8%) | 93 (36.0%) | 134 (36.0%) | 514 (26.1%) | 618 (26.6%) | 326 (42.8%) | 740 (29.6%) |
| **Former smoker** | 6 (0.7%) | 25 (0.9%) | <5 | <5 | 12 (0.6%) | 12 (0.5%) | <5 | 19 (0.8%) |
| **Current smoker** | 431 (52.7%) | 1502 (53.7%) | 76 (29.5%) | 119 (32.0%) | 815 (41.3%) | 1009 (43.5%) | 158 (20.8%) | 933 (37.3%) |
| **Missing** | 232 (28.4%) | 829 (29.6%) | 87 (33.7%) | 116 (31.2%) | 632 (32.0%) | 682 (29.4%) | 275 (36.1%) | 807 (32.3%) |
| **General conditions (any time prior)** | | | | | | | | |
| **Cerebrovascular disease** | 71 (8.7%) | 185 (6.6%) | 19 (7.4%) | 16 (4.3%) | 113 (5.7%) | 106 (4.6%) | 36 (4.7%) | 149 (6.0%) |
| **Chronic liver disease** | 15 (1.8%) | 36 (1.3%) | <5 | 5 (1.3%) | 25 (1.3%) | 39 (1.7%) | <5 | 42 (1.7%) |
| **Chronic obstructive lung disease** | 120 (14.7%) | 399 (14.3%) | 12 (4.7%) | 19 (5.1%) | 151 (7.7%) | 198 (8.5%) | 45 (5.9%) | 193 (7.7%) |
| **Coronary arteriosclerosis** | 9 (1.1%) | 43 (1.5%) | <5 | <5 | 14 (0.7%) | 19 (0.8%) | 5 (0.7%) | 16 (0.6%) |
| **Dementia** | 12 (1.5%) | 35 (1.3%) | <5 | <5 | 47 (2.4%) | 23 (1.0%) | 11 (1.4%) | 22 (0.9%) |
| **Depressive disorder** | 99 (12.1%) | 362 (12.9%) | 38 (14.7%) | 44 (11.8%) | 276 (14.0%) | 337 (14.5%) | 104 (13.7%) | 341 (13.6%) |
| **Diabetes mellitus** | 81 (9.9%) | 259 (9.3%) | 21 (8.1%) | 39 (10.5%) | 158 (8.0%) | 166 (7.2%) | 82 (10.8%) | 227 (9.1%) |
| **Gastroesophageal reflux disease** | 20 (2.4%) | 91 (3.3%) | 8 (3.1%) | <5 | 45 (2.3%) | 65 (2.8%) | 16 (2.1%) | 64 (2.6%) |
| **Gastrointestinal haemorrhage** | 45 (5.5%) | 178 (6.4%) | 9 (3.5%) | 26 (7.0%) | 119 (6.0%) | 143 (6.2%) | 43 (5.7%) | 162 (6.5%) |
| **Heart disease** | 142 (17.4%) | 490 (17.5%) | 41 (15.9%) | 48 (12.9%) | 293 (14.9%) | 293 (12.6%) | 131 (17.2%) | 297 (11.9%) |
| **Hyperlipidemia** | 89 (10.9%) | 251 (9.0%) | 17 (6.6%) | 31 (8.3%) | 147 (7.5%) | 170 (7.3%) | 67 (8.8%) | 193 (7.7%) |
| **Hypertensive disorder** | 232 (28.4%) | 701 (25.1%) | 79 (30.6%) | 73 (19.6%) | 482 (24.4%) | 495 (21.3%) | 185 (24.3%) | 564 (22.6%) |
| **Ischemic heart disease** | 75 (9.2%) | 285 (10.2%) | 27 (10.5%) | 28 (7.5%) | 161 (8.2%) | 149 (6.4%) | 69 (9.1%) | 152 (6.1%) |
| **Osteoarthritis** | 149 (18.2%) | 459 (16.4%) | 50 (19.4%) | 39 (10.5%) | 346 (17.5%) | 297 (12.8%) | 123 (16.2%) | 357 (14.3%) |
| **Peripheral vascular disease** | 39 (4.8%) | 105 (3.8%) | <5 | 7 (1.9%) | 58 (2.9%) | 53 (2.3%) | 14 (1.8%) | 65 (2.6%) |
| **Renal impairment** | 68 (8.3%) | 267 (9.5%) | 25 (9.7%) | 32 (8.6%) | 178 (9.0%) | 124 (5.3%) | 88 (11.6%) | 196 (7.8%) |
| **Venous thrombosis** | 34 (4.2%) | 95 (3.4%) | 11 (4.3%) | 15 (4.0%) | 80 (4.1%) | 67 (2.9%) | 29 (3.8%) | 85 (3.4%) |

### **Table 5: Overall incidence rates (per 100 000 person years) for head and neck subsites for whole population and stratified by sex and database.**

|  |  | **GOLD** | | **Aurum** | |
| --- | --- | --- | --- | --- | --- |
| **Cancer** | **Sex** | **Incidence**  **(100 000 pys)** | **Events (n)** | **Incidence**  **(100 000 pys)** | **Events (n)** |
| **Hypopharynx** | **Both** | 0.93 (0.87 to 1.00) | 818 | 0.61 (0.58 to 0.65) | 1,081 |
|  | **Female** | 0.43 (0.37 to 0.49) | 191 | 0.28 (0.24 to 0.31) | 245 |
|  | **Male** | 1.45 (1.34 to 1.57) | 627 | 0.95 (0.89 to 1.02) | 836 |
| **Larynx** | **Both** | 3.19 (3.07 to 3.31) | 2,797 | 3.10 (3.01 to 3.18) | 5,453 |
|  | **Female** | 1.20 (1.10 to 1.30) | 534 | 1.21 (1.14 to 1.29) | 1,071 |
|  | **Male** | 5.25 (5.03 to 5.47) | 2,263 | 4.99 (4.84 to 5.14) | 4,382 |
| **Nasal Cavity & Sinus** | **Both** | 0.29 (0.26 to 0.33) | 258 | 0.28 (0.25 to 0.30) | 488 |
|  | **Female** | 0.22 (0.18 to 0.27) | 98 | 0.22 (0.19 to 0.25) | 194 |
|  | **Male** | 0.37 (0.32 to 0.43) | 160 | 0.33 (0.30 to 0.38) | 294 |
| **Nasopharynx** | **Both** | 0.42 (0.38 to 0.47) | 372 | 0.45 (0.42 to 0.48) | 793 |
|  | **Female** | 0.27 (0.22 to 0.32) | 119 | 0.29 (0.25 to 0.33) | 255 |
|  | **Male** | 0.59 (0.52 to 0.66) | 253 | 0.61 (0.56 to 0.67) | 538 |
| **Oral Cavity** | **Both** | 2.25 (2.15 to 2.35) | 1,973 | 1.47 (1.41 to 1.52) | 2,584 |
|  | **Female** | 1.87 (1.74 to 2.00) | 833 | 1.18 (1.11 to 1.26) | 1,045 |
|  | **Male** | 2.64 (2.49 to 2.80) | 1,140 | 1.75 (1.67 to 1.84) | 1,539 |
| **Oropharynx** | **Both** | 2.65 (2.54 to 2.76) | 2,321 | 2.46 (2.39 to 2.54) | 4,340 |
|  | **Female** | 1.35 (1.24 to 1.46) | 600 | 1.33 (1.25 to 1.41) | 1,173 |
|  | **Male** | 3.99 (3.80 to 4.18) | 1,721 | 3.61 (3.48 to 3.73) | 3,167 |
| **Salivary Gland** | **Both** | 0.87 (0.81 to 0.93) | 761 | 0.89 (0.85 to 0.94) | 1,573 |
|  | **Female** | 0.70 (0.62 to 0.78) | 310 | 0.80 (0.74 to 0.86) | 706 |
|  | **Male** | 1.05 (0.95 to 1.15) | 451 | 0.99 (0.92 to 1.05) | 867 |
| **Tongue** | **Both** | 2.85 (2.74 to 2.96) | 2,499 | 2.51 (2.43 to 2.58) | 4,418 |
|  | **Female** | 1.91 (1.78 to 2.04) | 849 | 1.67 (1.59 to 1.76) | 1,475 |
|  | **Male** | 3.82 (3.64 to 4.01) | 1,650 | 3.35 (3.23 to 3.47) | 2,943 |

### **Figure 1: Annualised incidence rates from 2000 to 2021 for HNC and subsites stratified by database and sex**

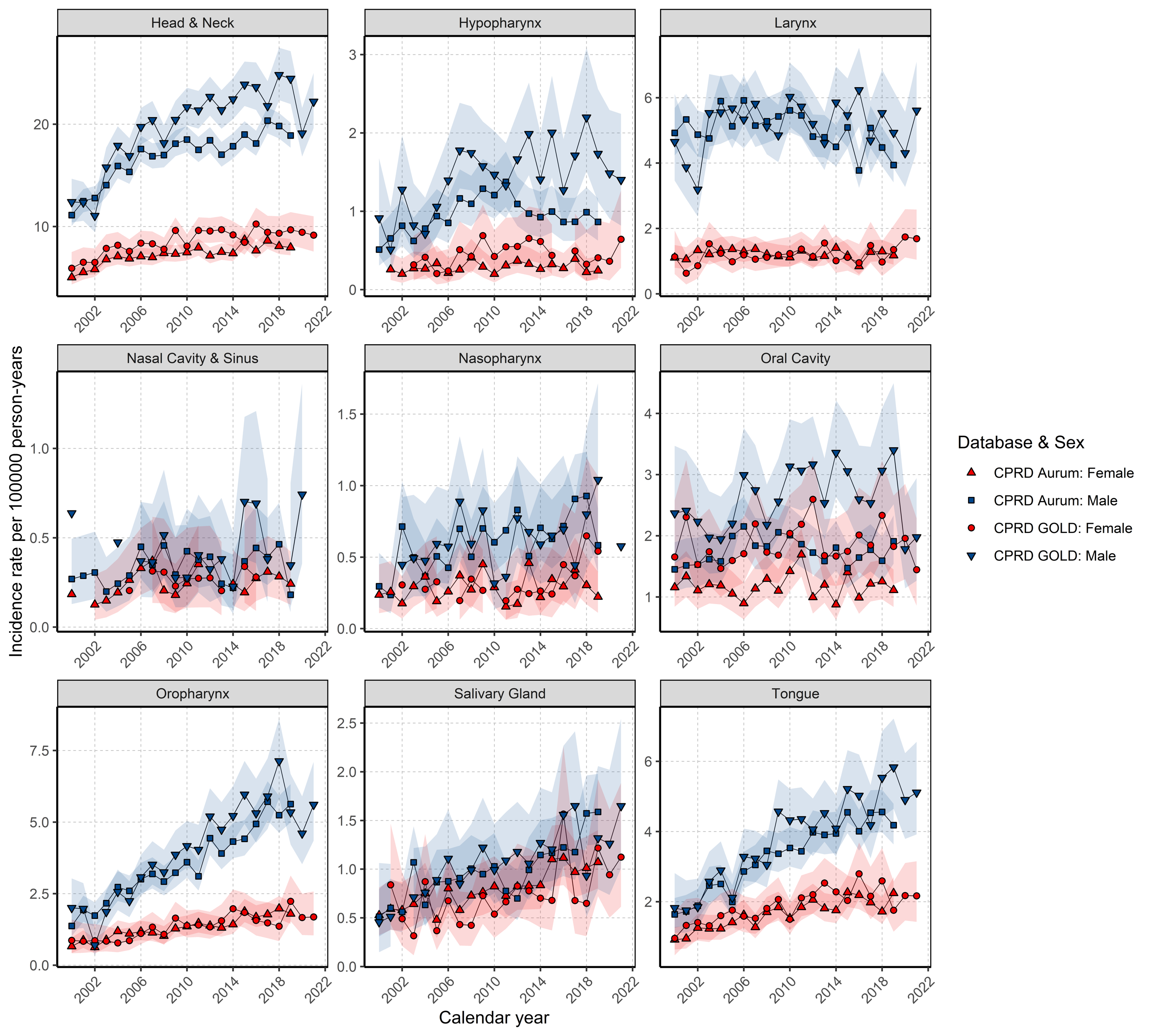

### **Figure 2: Age standardized Annualised incidence rates from 2000 to 2021 for HNC and subsites stratified by database and sex using the European Standard Population 2013 in CPRD GOLD. Red line depicting when Quality and Outcomes Framework (QOF) was introduced in the UK in 2004.**

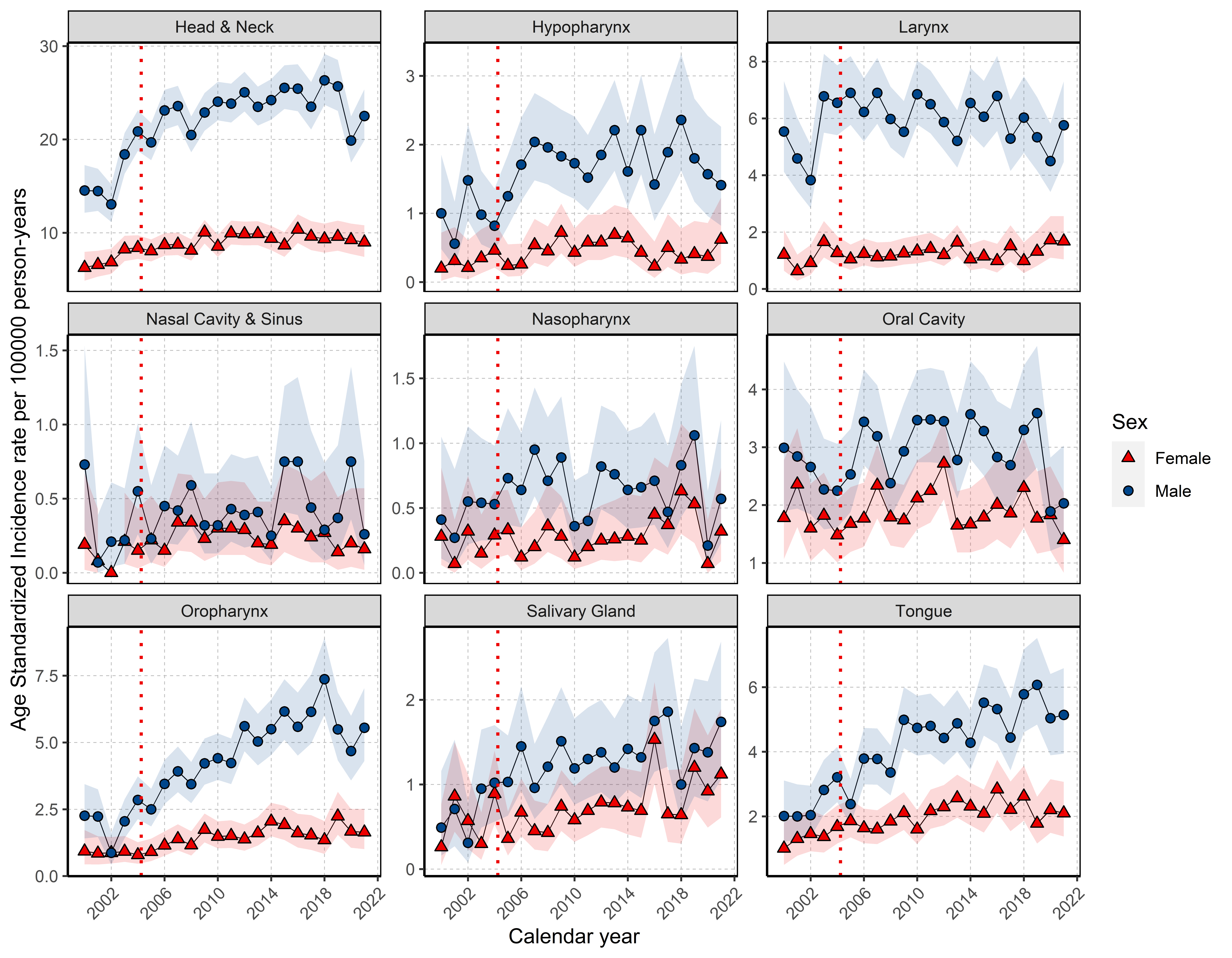

### **Figure 3: Age standardized Annualised incidence rates from 2002 to 2020 for HNC compared with national cancer registries across the UK using the European Standard Population 2013.**

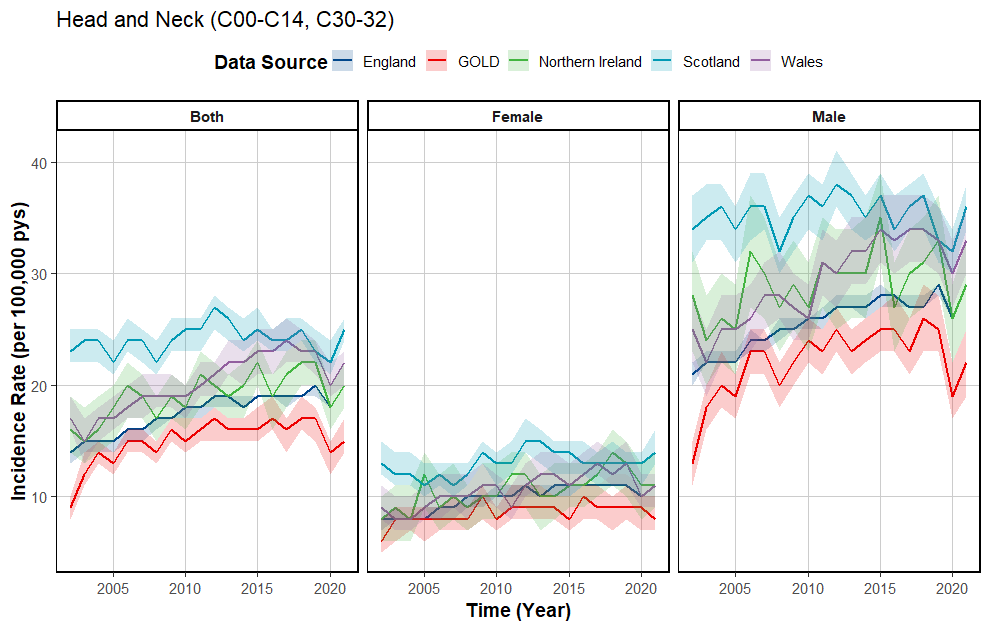

### **Figure 4: Overall IR from 2000 to 2021 for HNC and subsites per age group stratified by database.**

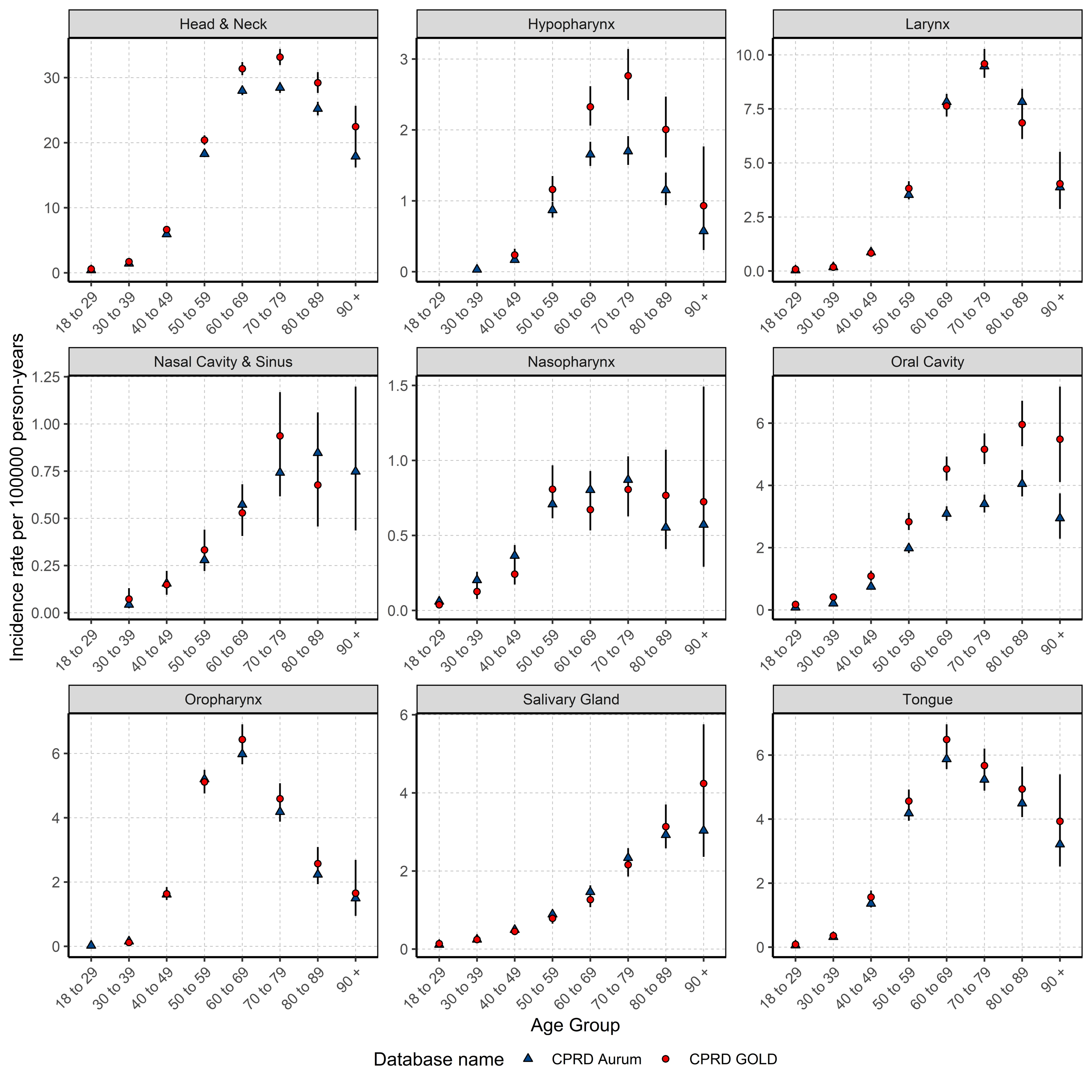

### **Figure 5: Annualised incidence rates for all HNC stratified by database and age group.**

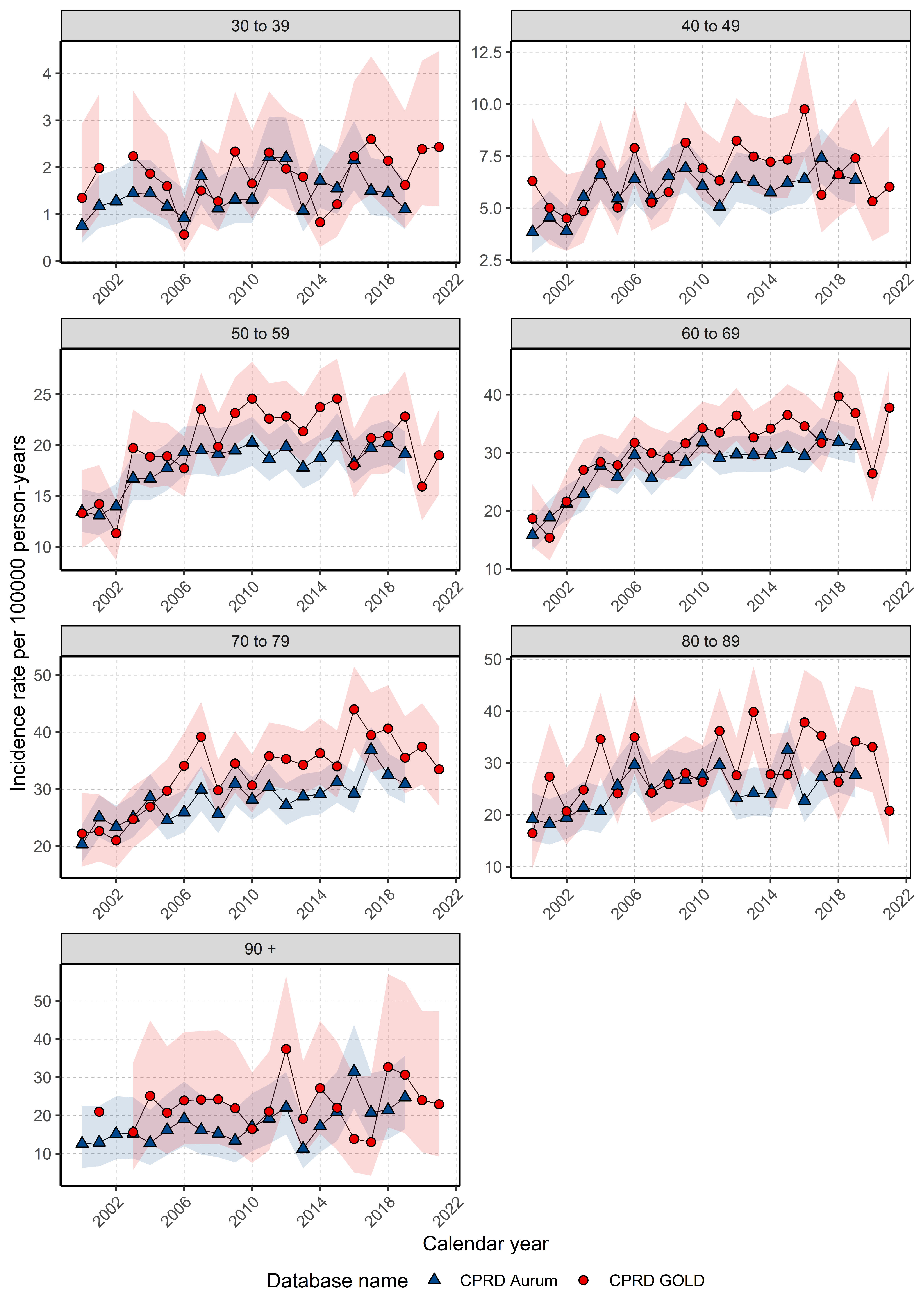

### **Figure 6: Annualised incidence rates for all HNC stratified by database, sex and age group.**

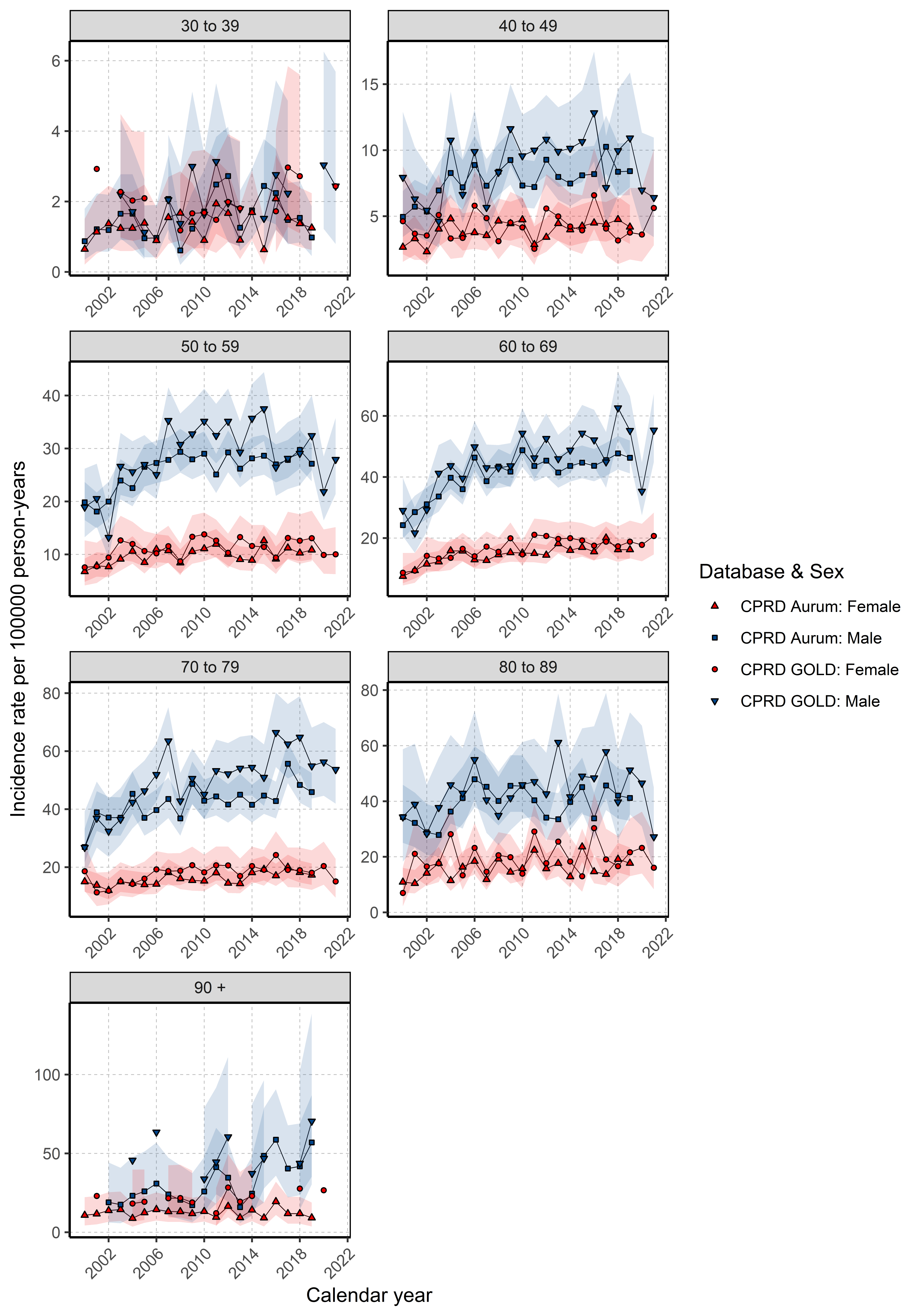

### **Figure 7: Annualised IR for all HNC subsites from 2000 to 2021 stratified by database, and age group.**

#### **Figure 7.1: Annualised IR from 2000 to 2021 stratified by database and age group for cancer of the hypopharynx.**

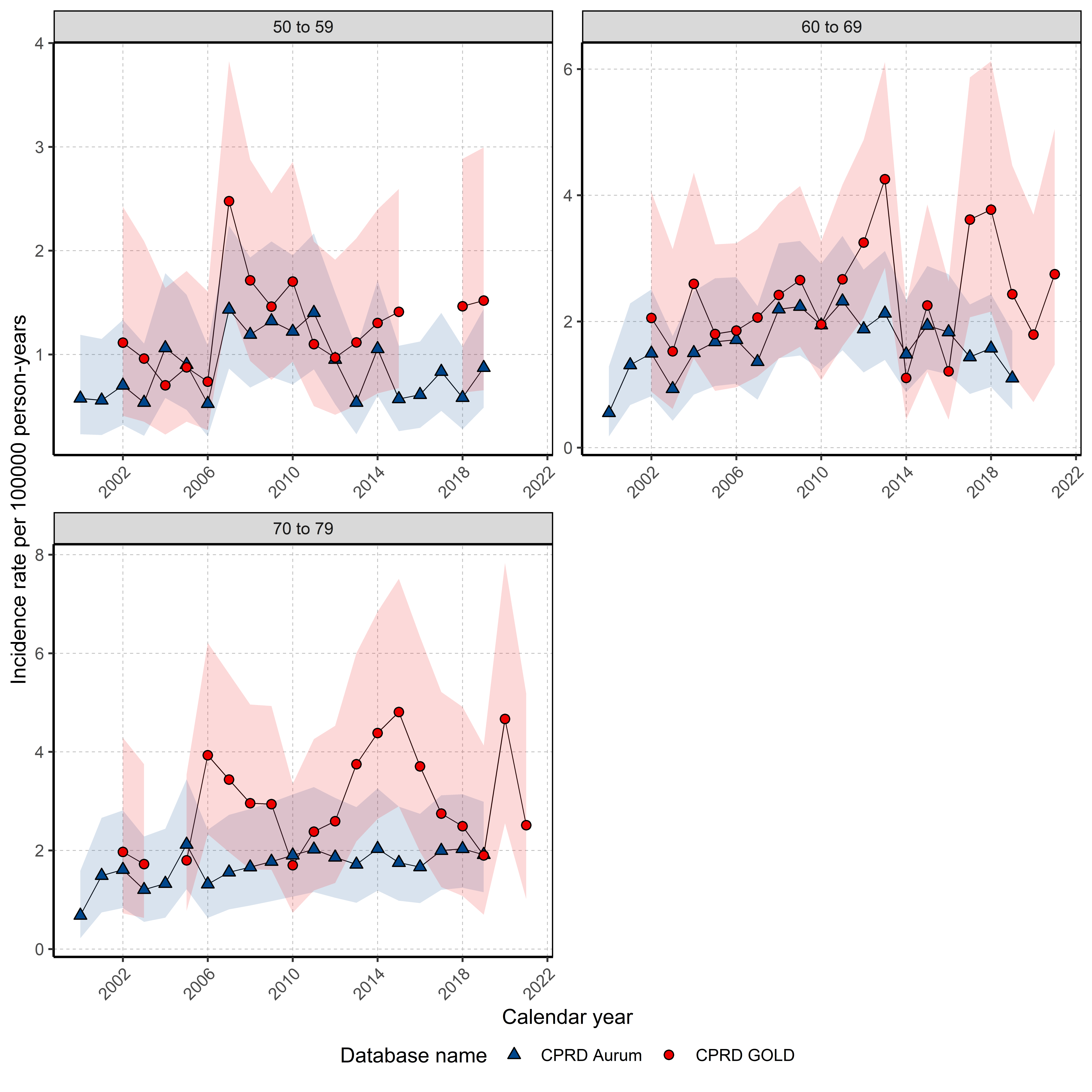

#### **Figure 7.2: Annualised IR from 2000 to 2021 stratified by database and age group for cancer of the larynx.**

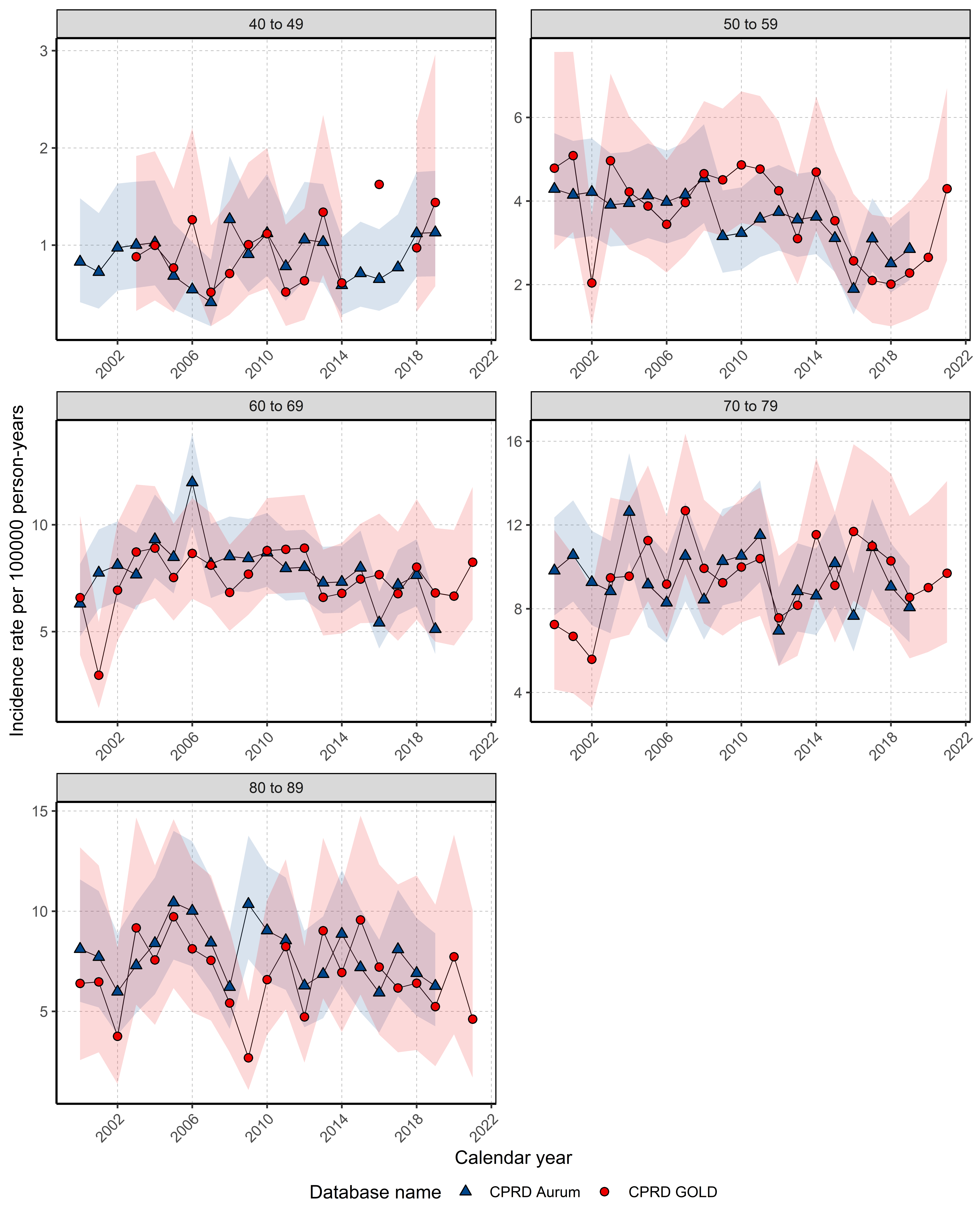

#### **Figure 7.3: Annualised IR from 2000 to 2021 stratified by database and age group for cancer of the oral cavity.**

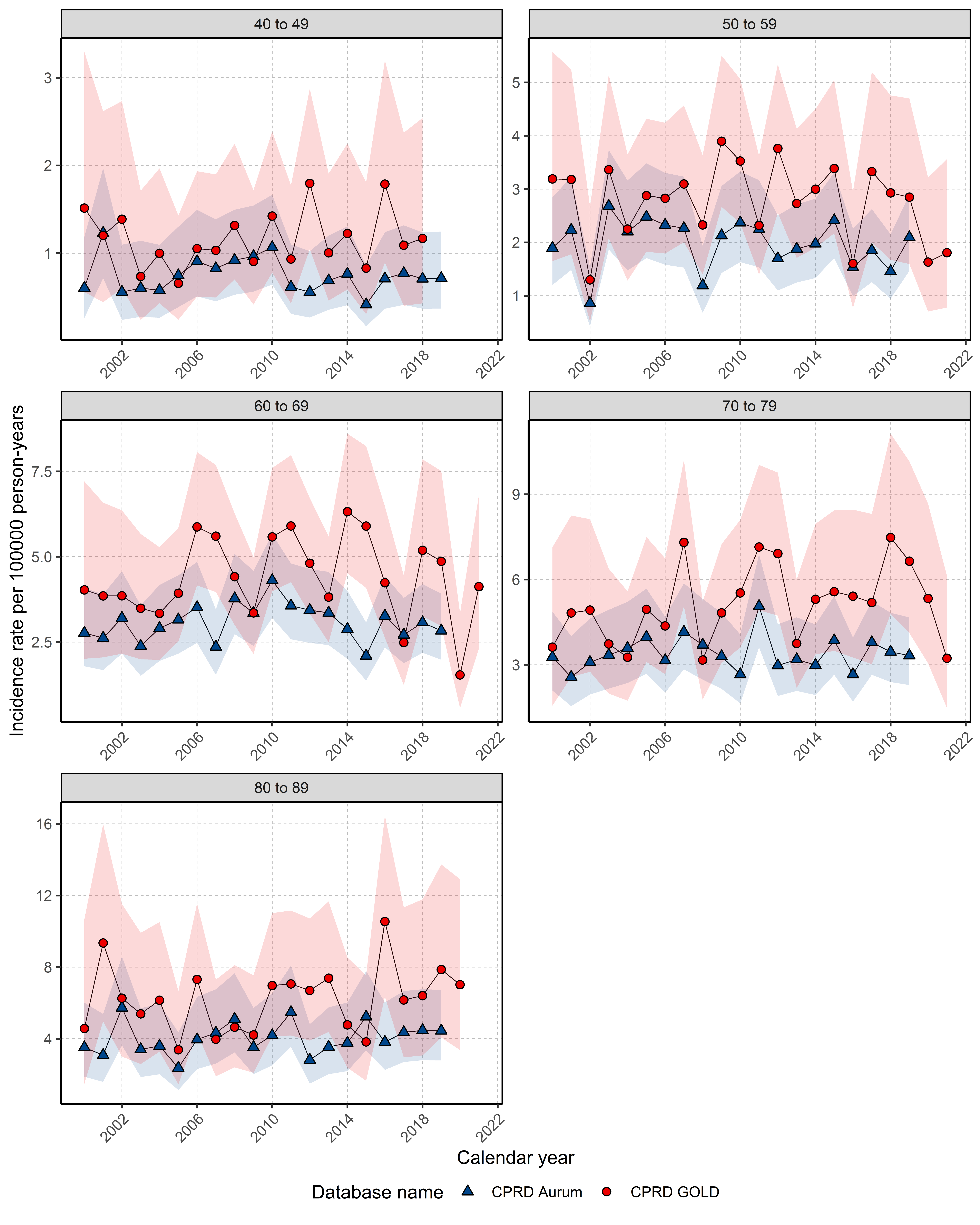

#### **Figure 7.4: Annualised IR from 2000 to 2021 stratified by database and age group for cancer of the oropharynx.**

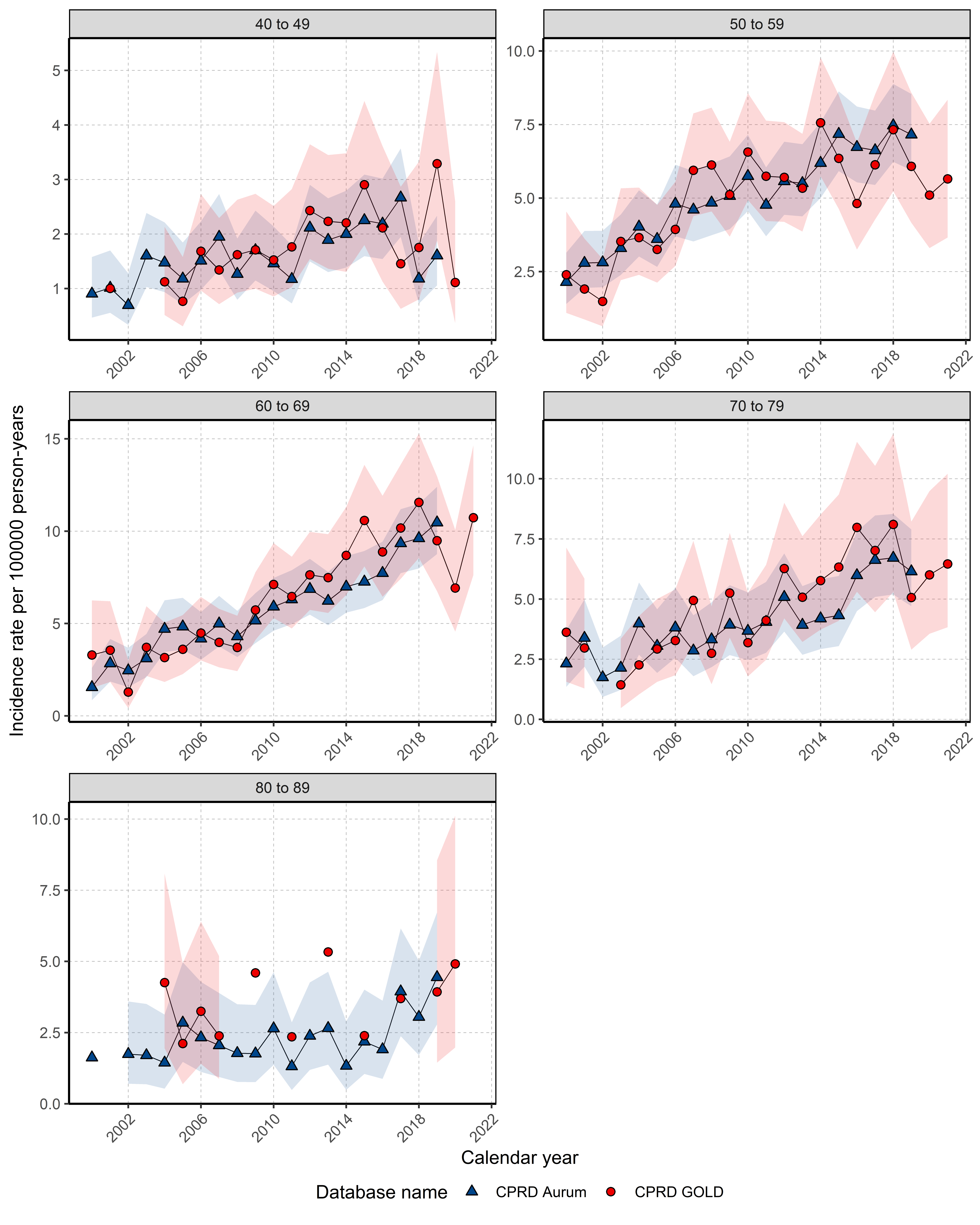

#### **Figure 7.5: Annualised IR from 2000 to 2021 stratified by database and age group for cancer of the salivary gland.**

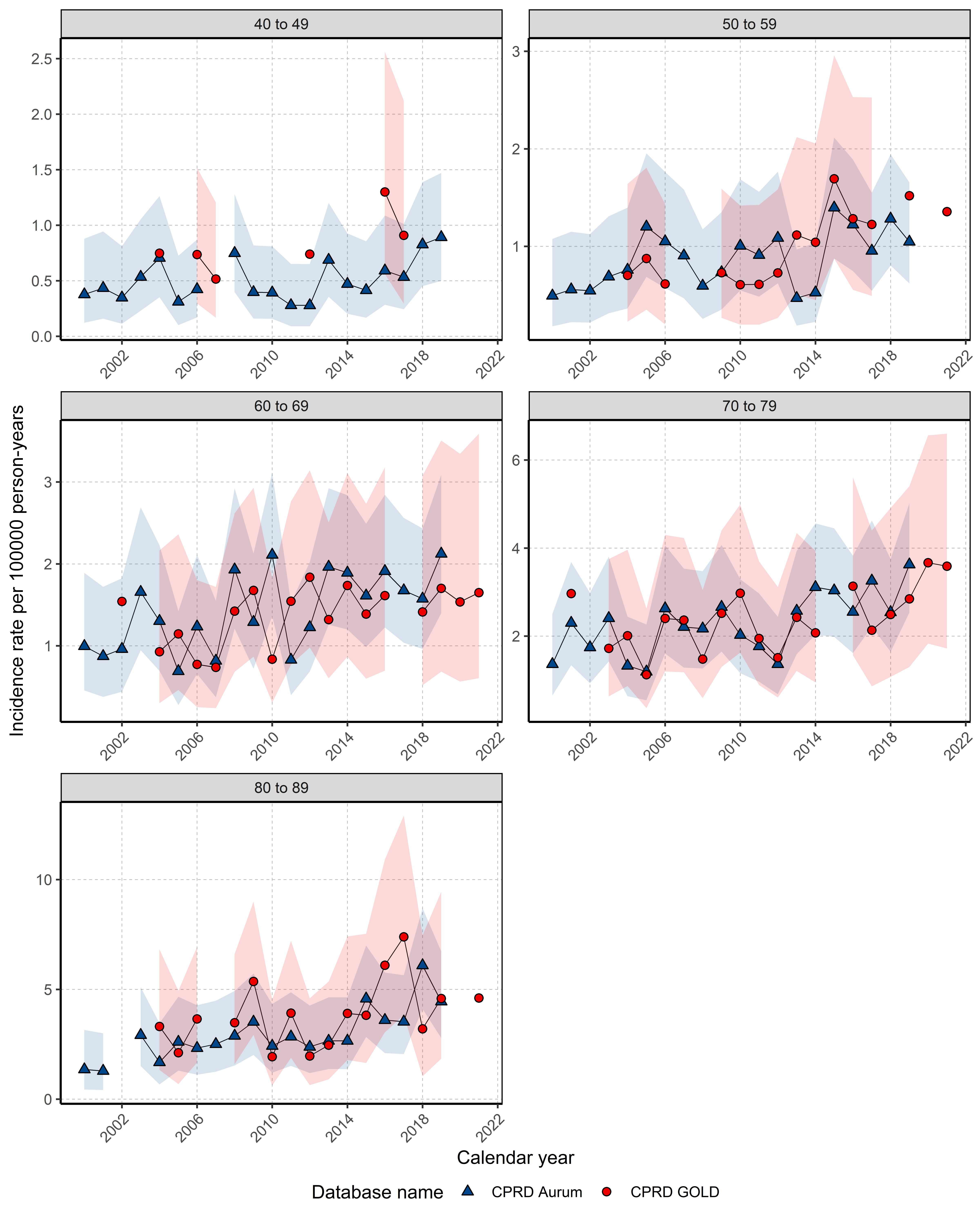

#### **Figure 7.6: Annualised IR from 2000 to 2021 stratified by database and age group for cancer of the tongue.**

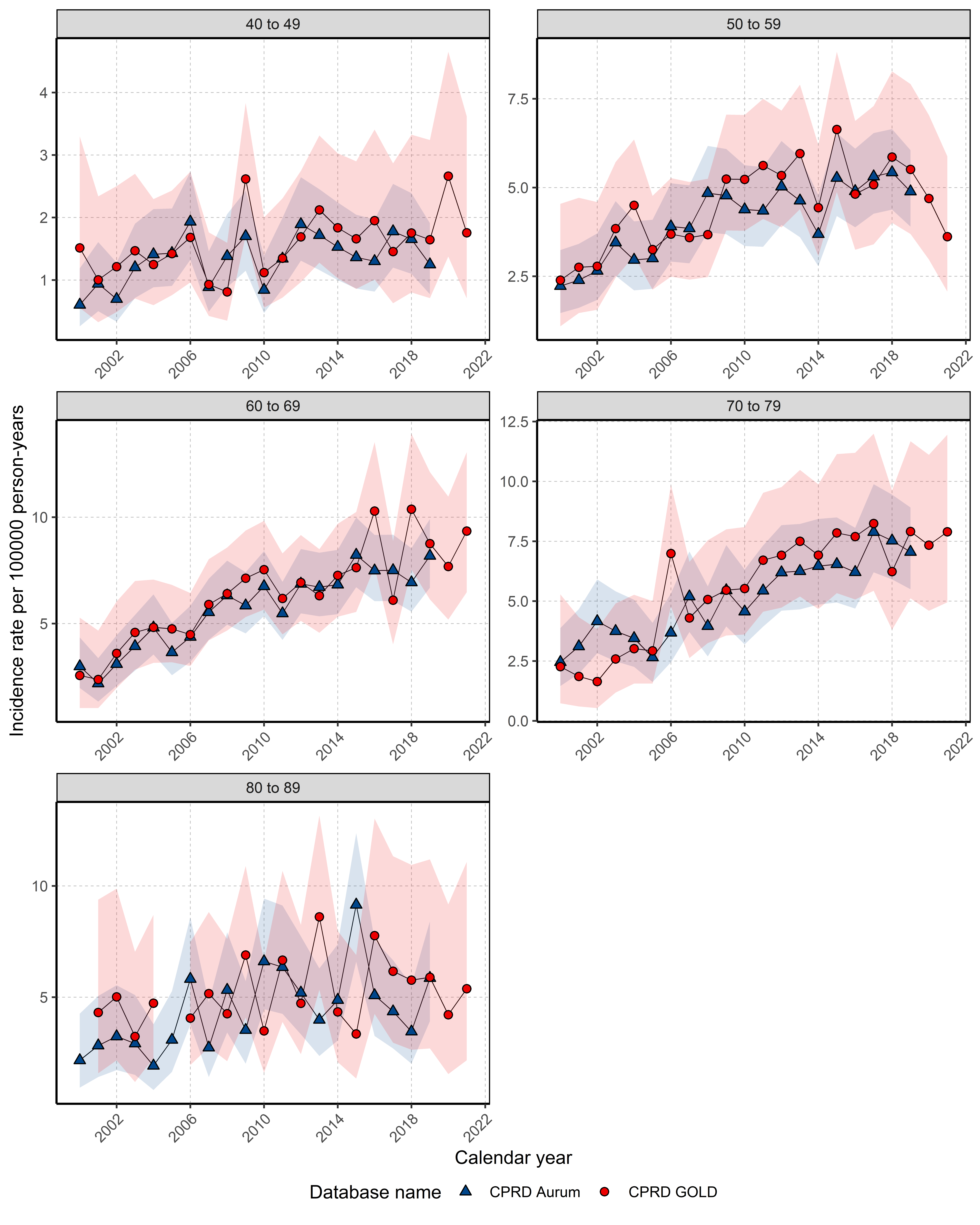

### **Figure 8: Kaplan-Meier survival curves of HNC and subsites by database**

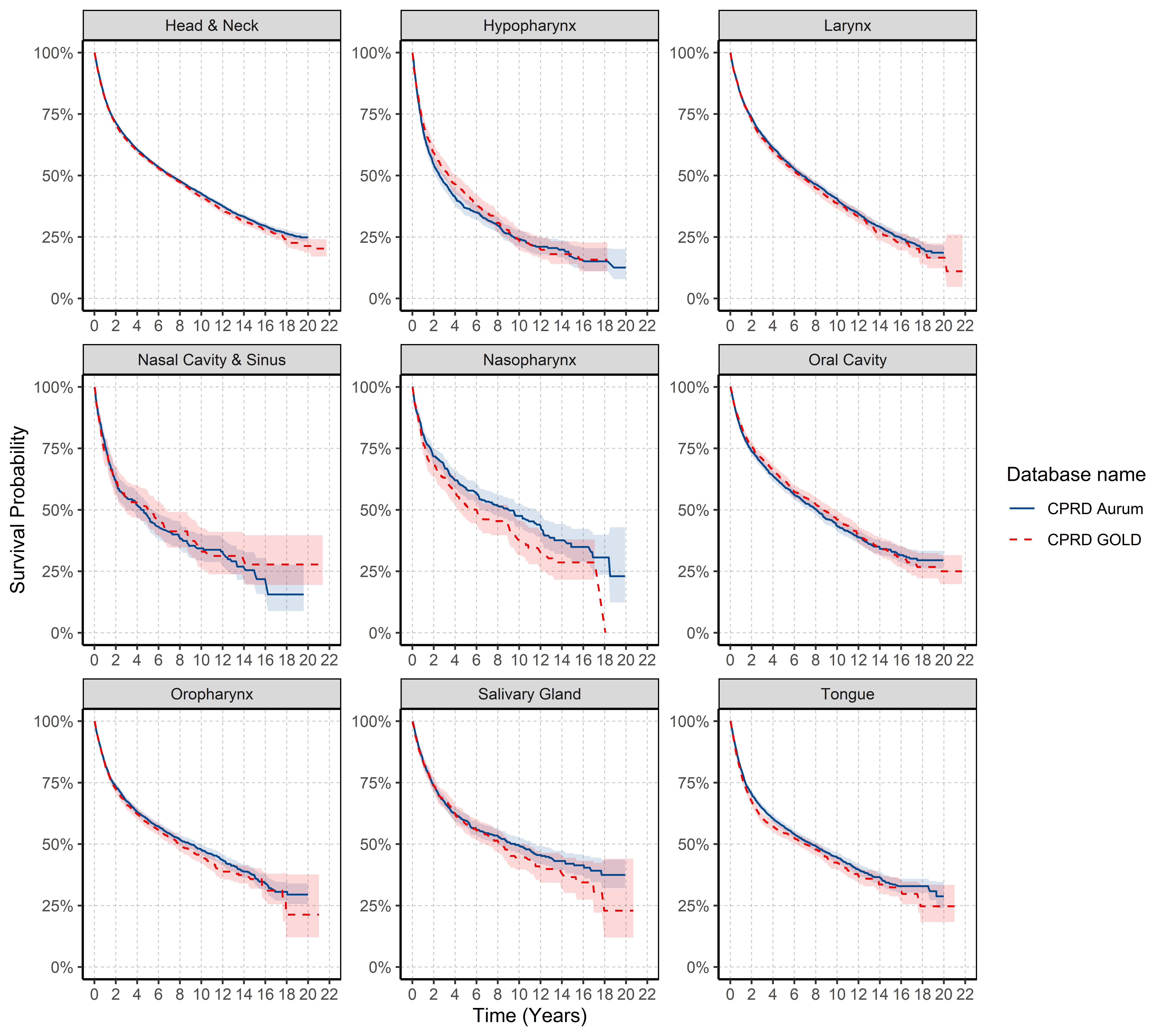

### **Table 6: Median survival in years for HNC and subsites by database.**

| **Cancer** | **Database** | **Records (n)** | **Events (n)** | **Median Survival in Years (95% CI)** |
| --- | --- | --- | --- | --- |
| Hypopharynx | Aurum | 1068 | 676 | 2.560 (2.155 - 3.075) |
|  | GOLD | 808 | 481 | 3.373 (2.867 - 4.096) |
| Larynx | Aurum | 5408 | 2724 | 6.757 (6.319 - 7.255) |
|  | GOLD | 2768 | 1359 | 6.535 (5.971 - 7.211) |
| Nasal Cavity & Sinus | Aurum | 479 | 265 | 4.405 (3.146 - 5.618) |
|  | GOLD | 252 | 133 | 5.054 (2.825 - 6.702) |
| Nasopharynx | Aurum | 791 | 322 | 9.112 (6.505 - 11.661) |
|  | GOLD | 368 | 177 | 6.056 (4.309 - 9.060) |
| Oral Cavity | Aurum | 2574 | 1161 | 8.041 (7.368 - 8.690) |
|  | GOLD | 1962 | 820 | 8.674 (7.959 - 9.837) |
| Oropharynx | Aurum | 4319 | 1694 | 8.986 (7.992 - 9.936) |
|  | GOLD | 2307 | 937 | 7.984 (7.192 - 9.027) |
| Salivary Gland | Aurum | 1569 | 610 | 9.621 (8.060 - 11.373) |
|  | GOLD | 756 | 303 | 8.435 (6.762 - 10.836) |
| Tongue | Aurum | 4404 | 1891 | 7.603 (6.883 - 8.372) |
|  | GOLD | 2484 | 1110 | 7.006 (6.053 - 8.162) |

### **Figure 9: Kaplan-Meier survival curve of HNC and subtypes for CPRD GOLD stratified by calendar year of diagnosis (2000-2004, 2005-2009, 2010-2014, 2015-2019 and 2020-2021)**

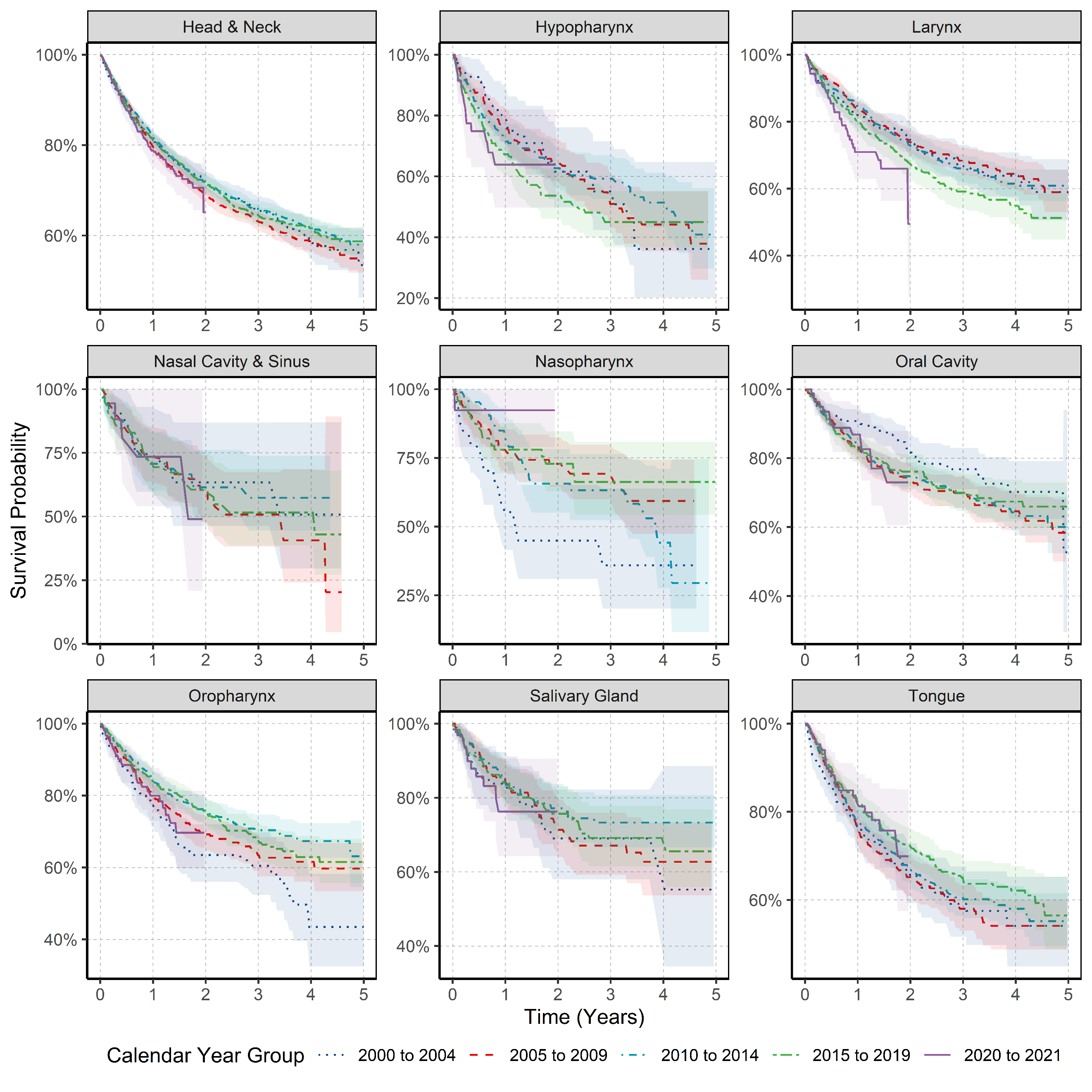
